# Next-generation phenotyping integrated in a national framework for patients with ultra-rare disorders improves genetic diagnostics and yields new molecular findings

**DOI:** 10.1101/2023.04.19.23288824

**Authors:** Axel Schmidt, Magdalena Danyel, Kathrin Grundmann, Theresa Brunet, Hannah Klinkhammer, Tzung-Chien Hsieh, Hartmut Engels, Sophia Peters, Alexej Knaus, Shahida Moosa, Luisa Averdunk, Felix Boschann, Henrike Sczakiel, Sarina Schwartzmann, Martin Atta Mensah, Jean Tori Pantel, Manuel Holtgrewe, Annemarie Bösch, Claudia Weiß, Natalie Weinhold, Aude-Annick Suter, Corinna Stoltenburg, Julia Neugebauer, Tillmann Kallinich, Angela M. Kaindl, Susanne Holzhauer, Christoph Bührer, Philip Bufler, Uwe Kornak, Claus-Eric Ott, Markus Schülke, Hoa Huu Phuc Nguyen, Sabine Hoffjan, Corinna Grasemann, Tobias Rothoeft, Folke Brinkmann, Nora Matar, Sugirthan Sivalingam, Claudia Perne, Elisabeth Mangold, Martina Kreiss, Kirsten Cremer, Regina C. Betz, Tim Bender, Martin Mücke, Lorenz Grigull, Thomas Klockgether, Spier Isabel, Heimbach André, Bender Tim, Fabian Brand, Christiane Stieber, Alexandra Marzena Morawiec, Pantelis Karakostas, Valentin S. Schäfer, Sarah Bernsen, Patrick Weydt, Sergio Castro-Gomez, Ahmad Aziz, Marcus Grobe-Einsler, Okka Kimmich, Xenia Kobeleva, Demet Önder, Hellen Lesmann, Sheetal Kumar, Pawel Tacik, Min Ae Lee-Kirsch, Reinhard Berner, Catharina Schuetz, Julia Körholz, Tanita Kretschmer, Nataliya Di Donato, Evelin Schröck, André Heinen, Ulrike Reuner, Amalia-Mihaela Hanßke, Frank J. Kaiser, Eva Manka, Martin Munteanu, Alma Kuechler, Kiewert Cordula, Raphael Hirtz, Elena Schlapakow, Christian Schlein, Jasmin Lisfeld, Christian Kubisch, Theresia Herget, Maja Hempel, Christina Weiler-Normann, Kurt Ullrich, Christoph Schramm, Cornelia Rudolph, Franziska Rillig, Maximilian Groffmann, Ania Muntau, Alexandra Tibelius, Eva M. C. Schwaibold, Christian P. Schaaf, Michal Zawada, Lilian Kaufmann, Katrin Hinderhofer, Pamela M. Okun, Urania Kotzaeridou, Georg F. Hoffmann, Daniela Choukair, Markus Bettendorf, Malte Spielmann, Annekatrin Ripke, Martje Pauly, Alexander Münchau, Katja Lohmann, Irina Hüning, Britta Hanker, Tobias Bäumer, Rebecca Herzog, Yorck Hellenbroich, Dominik S. Westphal, Tim Strom, Reka Kovacs, Korbinian M. Riedhammer, Katharina Mayerhanser, Elisabeth Graf, Melanie Brugger, Julia Hoefele, Konrad Oexle, Nazanin Mirza-Schreiber, Riccardo Berutti, Ulrich Schatz, Martin Krenn, Christine Makowski, Heike Weigand, Sebastian Schröder, Meino Rohlfs, Vill Katharina, Fabian Hauck, Ingo Borggraefe, Wolfgang Müller-Felber, Ingo Kurth, Miriam Elbracht, Cordula Knopp, Matthias Begemann, Florian Kraft, Johannes R. Lemke, Julia Hentschel, Konrad Platzer, Vincent Strehlow, Rami Abou Jamra, Martin Kehrer, German Demidov, Stefanie Beck-Wödl, Holm Graessner, Marc Sturm, Lena Zeltner, Ludger J. Schöls, Janine Magg, Andrea Bevot, Christiane Kehrer, Nadja Kaiser, Denise Horn, Annette Grüters-Kieslich, Christoph Klein, Stefan Mundlos, Markus Nöthen, Olaf Riess, Thomas Meitinger, Heiko Krude, Peter M. Krawitz, Tobias Haack, Nadja Ehmke, Matias Wagner

## Abstract

Most individuals with rare diseases initially consult their primary care physician. For a subset of rare diseases, efficient diagnostic pathways are available. However, ultra-rare diseases often require both expert clinical knowledge and comprehensive genetic diagnostics, which poses structural challenges for public healthcare systems. To address these challenges within Germany, a novel structured diagnostic concept, based on multidisciplinary expertise at established university hospital centers for rare diseases (CRDs), was evaluated in the three year prospective study TRANSLATE NAMSE. A key goal of TRANSLATE NAMSE was to assess the clinical value of exome sequencing (ES) in the ultra-rare disease population. The aims of the present study were to perform a systematic investigation of the phenotypic and molecular genetic data of TRANSLATE NAMSE patients who had undergone ES in order to determine the yield of both ultra-rare diagnoses and novel gene-disease associations; and determine whether the complementary use of machine learning and artificial intelligence (AI) tools improved diagnostic effectiveness and efficiency.

ES was performed for 1,577 patients (268 adult and 1,309 pediatric). Molecular genetic diagnoses were established in 499 patients (74 adult and 425 pediatric). A total of 370 distinct molecular genetic causes were established. The majority of these concerned known disorders, most of which were ultra-rare. During the diagnostic process, 34 novel and 23 candidate genotype-phenotype associations were delineated, mainly in individuals with neurodevelopmental disorders.

To determine the likelihood that ES will lead to a molecular diagnosis in a given patient, based on the respective clinical features only, we developed a statistical framework called YieldPred. The genetic data of a subcohort of 224 individuals that also gave consent to the computer-assisted analysis of their facial images were processed with the AI tool Prioritization of Exome Data by Image Analysis (PEDIA) and showed superior performance in variant prioritization.

The present analyses demonstrated that the novel structured diagnostic concept facilitated the identification of ultra-rare genetic disorders and novel gene-disease associations on a national level and that the machine learning and AI tools improved diagnostic effectiveness and efficiency for ultra-rare genetic disorders.

## Introduction

A recent analysis of the Orphanet database showed that around 3% to 6% of the global population have a rare disease (i.e., a disease with a prevalence of < 1 in 2,000), and that 72% of such cases may have a genetic cause^1^. Rare diseases thus represent a substantial global health burden. However, only a minority of suspected rare disease patients receive both a definite clinical diagnosis and a confirmatory molecular test result^2,3^. The International Rare Disease Research Consortium therefore stated that by 2027, all patients who come to medical attention with a suspected rare disease should be diagnosed within one year, if the respective disorder has been described in the medical literature^4^. Since many rare diseases are Mendelian in nature, comprehensive genetic testing is a key element to achieve that goal.

In Germany, around 90% of the population has statutory health insurance, and the current reimbursement scheme allows physicians to request chromosome analyses, molecular karyotyping, and the sequencing of single genes or gene panels. For example, high-resolution genome-wide array-based segmental aneusomy profiling detects a pathogenic aberration in around 19% of patients with developmental delay^5^. Besides contiguous gene syndromes, most of the remaining rare disorders are monogenic, and are caused by single nucleotide variants or small insertions or deletions (indels). However, single gene analyses or small gene panels are only likely to detect a pathogenic aberration if the phenotype is highly predictive of the molecular cause e.g., hemoglobinopathies^6^.

For phenotypes with high genetic heterogeneity, such as neurodevelopmental disorders, genetic investigation is more challenging. For intellectual disability, for example, studies to date have identified disease associations for more than a thousand genes^7^. For these disorders, research has shown that exome sequencing (ES) can be more cost-effective than gene panel sequencing^8^. However, this is also accompanied by more genetic variants that have to be assessed. Therefore, a clear indication for ES and efficient data analysis strategies are crucial. Between 2018 and 2020, a novel diagnostic concept within the German healthcare system was evaluated in the prospective study TRANSLATE NAMSE^9^.

This involved standardized structures and procedures and multidisciplinary teams (MDTs) at 10 university hospital-based Centers for Rare Diseases (CRDs). The MDTs conducted a three step diagnostic process: 1) primary review of patient records; 2) selection of diagnostic procedures, including a possible recommendation for ES; and 3) evaluation of all findings, including genetic variants. A key goal was to investigate whether ES would facilitate the diagnosis of ultra-rare disorders, or even the delineation of novel monogenic disorders. In this work we report about the molecular findings of this study.

Furthermore, in a companion study, we also investigated to which extent the results from computer-assisted pattern recognition in facial dysmorphism contribute to variant interpretation (PEDIA, Prioritization of Exome Data by Image Analysis) and how phenotypic features can be used to estimate the probability that a molecular diagnosis can be established with ES (YieldPred). The present analyses demonstrated that ES facilitated the diagnosis of ultra-rare genetic diseases and novel gene-disease associations, and that AI-driven technologies improved diagnostic effectiveness and efficiency for ultra-rare genetic disorders.

## Results

### Phenotypic characteristics of the study cohort

Between 2018 and 2020, a total of 5,652 individuals (2,033 adults, 3,619 children) with a suspected rare disorder were enrolled in TRANSLATE NAMSE by CRDs at 10 German university hospitals (Figure 1a)^9^. The present analyses were performed using the data of a total of 1,577 of these 5,652 patients (268 adults and 1,309 children), i.e., those individuals who had been referred for ES on the recommendation of the MDT at the respective CRD (ES cohort, Supplemental Table 1).

**Figure 1:**
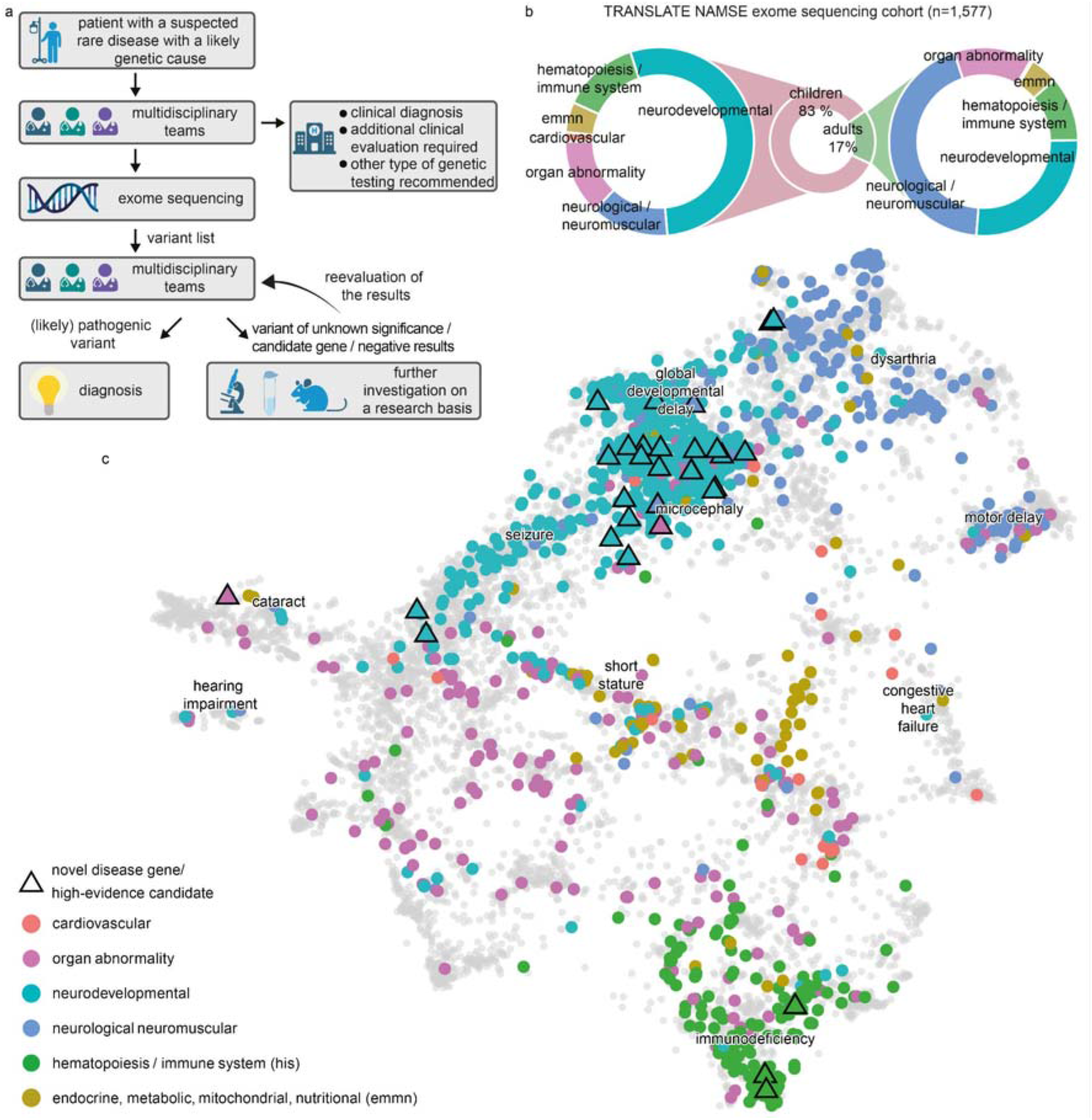
Workflow in the TRANSLATE NAMSE project and phenotypes in which exome sequencing was performed. a) Patients with a suspected rare disease were referred to a multidisciplinary team (MDT) and deeply phenotyped using Human Phenotype Ontology (HPO) terminology. If a genetic etiology was considered likely, exome sequencing (ES) was performed. The MDT then evaluated the molecular findings, and could order additional analyses for variants of unknown significance or variants in potentially novel disease candidate genes (created with BioRender.com). b) ES was performed predominantly in children. The main indications for ES in children were neurodevelopmental disorders. In adults, the main indications were neurological/neuromuscular disorders. In both children and adults, the least common disease categories were cardiovascular, <u>e</u>ndocrine, <u>m</u>etabolic, <u>m</u>itochondrial, <u>n</u>utritional (emmn), and hematopoiesis/immune system (his). c) Phenotypic similarities between patients, as encoded according to their HPO terms, were visualized with Uniform Manifold Approximation and Projection (UMAP). The clinical phenotype space was initially defined by all OMIM diseases, using their HPO annotations (gray background dots). For each patient, color-coding indicates allocation to disease groups, in accordance with the leading clinical feature. An overlap is evident for patients in the neurodevelopmental and neuromuscular groups (aquamarine and blue clusters), which indicates high phenotypic similarity. This precludes the unequivocal assignment of these patients to a diagnostic group. Triangles indicate patients who contributed to the identification of a novel, high evidence gene-phenotype association.

Each of these 1,577 individuals was assigned to one of six major disease categories by the respective CRD physician (Figure 1b). The majority of children were assigned to the disease category “neurodevelopmental disorders” (n=702, 54%), and the majority of adults were assigned to the disease category “neurological or neuromuscular disorders” (n=117, 44%). Smaller proportions of adult and pediatric cases were assigned to the groups “organ malformation”, “endocrine/metabolic disorders”, “immune/hematologic disorders”, and “cardiovascular disorders”. Patient phenotypes were also annotated as Human Phenotype Ontology (HPO) terms by the respective CRD physician. On average, five HPO terms were specified per individual (Supplemental Figure 1a). The phenotypes within the present cohort were visualized by projection into a clinical feature space, in which individuals were positioned according to their original HPO annotations. While most patients from the same disease group were in close proximity, the clusters showed a partial overlap (Figure 1c). For example, many patients categorized within “neurological or neuromuscular disorders” also showed HPO terms typically associated with “neurodevelopmental disorders” and vice versa (Supplemental Figure 1b).This suggests that grouping patients into single disease groups may be overly-simplistic.

### Diagnostic yield of ES

A molecular diagnosis was established in a total of 499 of the 1,577 patients (32%), i.e., in these cases, ES identified variants that fully or partially explained the phenotype. The diagnostic yield was slightly higher in children (32%) than in adults (28%, p=0.13, Fisher’s exact test, Figure 2a) and two-fold higher in patients assigned to the category “neurodevelopmental disorder” than for all other disease categories (42% vs. 22%, p<0.001, Fisher’s exact test with Bonferroni correction; for single comparisons between disorder groups see Figure 2b). Furthermore, ES found variants of unknown significance. Specifically, these variants were enriched for missense variants (80% vs. 45%, p<0.001, Supplemental Figure 2), due to lower support for pathogenicity according to the guidelines of the American College of Medical Genetics (ACMG) for interpretation of sequence variants.

**Figure 2:**
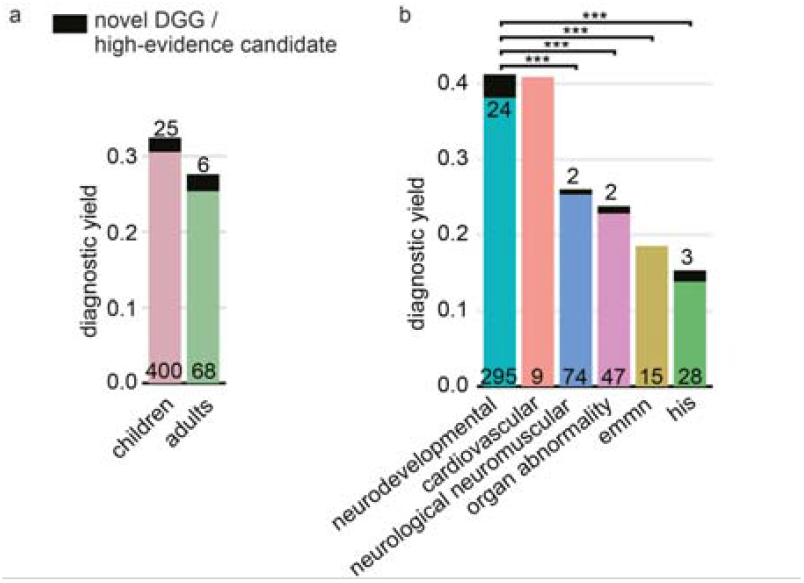
Diagnostic yield of exome sequencing depends on age and disease group. The diagnostic yield differed according to age group (adult/child) (a) and disease category (b). For all disease categories, with the exception of cardiovascular, the diagnostic yield was increased by novel diagnostic-grade genes (DGG) and high-evidence candidate genes (dark-colored tip of the bar). The absolute number of solved cases in which a variant was found in an established disease gene is given at the bottom of each bar, and the number of solved cases attributable to a novel DGG or high-evidence candidate gene is given at the top of each bar. The entire TRANSLATE NAMSE ES cohort was considered for a and b (n=1,577). Diagnostic yield between disease categories were compared using Fisher’s exact test. P-values were adjusted by Bonferroni correction. *** = p<0.001; emnn: endocrine, metabolic, mitochondrial, nutritional; his: hematopoiesis/immune system.

### *De novo* variants and parental mosaicism

A total of 228 diagnoses (45% of 510 diagnoses) were attributable to *de novo* variants (Figure 3a). In three families with variants that were initially classified as *de novo*, evidence for probable or certain parental mosaicism was found (Supplemental Material). In one of these families, the same likely pathogenic variant in *PUF60* was identified as the cause of developmental delay in two affected brothers. Since the variant was not detectable in the exome data of either parent, gonadal mosaicism could not be confirmed, and was instead presumed on the basis of the family history. The detection in the ES analysis of three probable parental mosaics among 228 patients, corresponds to a frequency of 1.3% which is within the estimated interval of clinically relevant parental mosaicism^10–12^.

**Figure 3:**
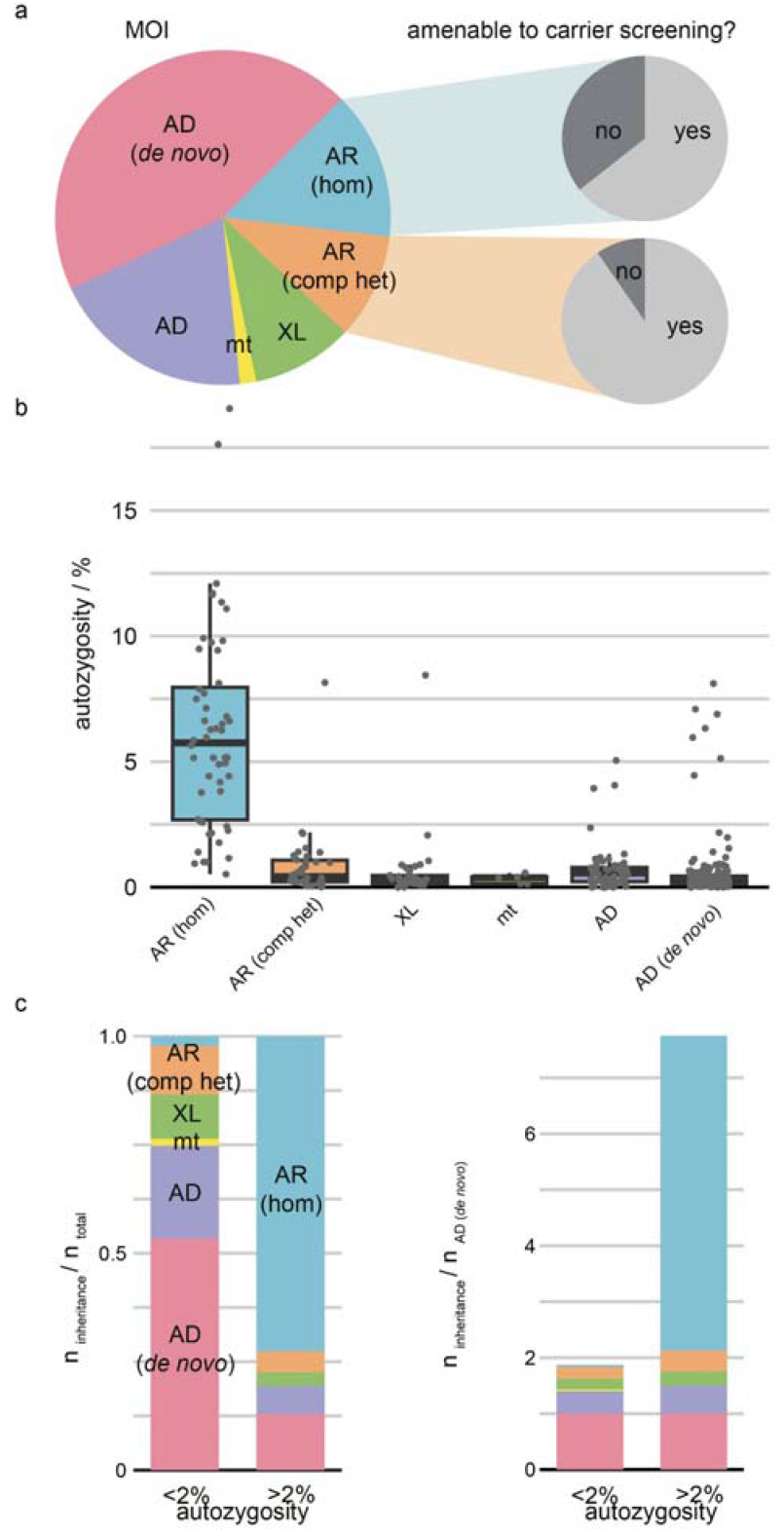
Mode of inheritance and disease burden are dependent on autozygosity. a) Pie chart showing the distribution of modes of inheritance (MOI) for all diagnoses (n=510). Most disease-causing variants occurred *de novo* and on an autosome. At least 75% of all autosomal recessive diagnoses could have been identified by expanded carrier screening (slice). b) Box plots of autozygosity for each MOI (n=375). Individuals are indicated by gray dots. Autozygosity was substantially increased in individuals with autosomal recessive disorders due to homozygous variants. (c) Bar graphs illustrating MOI in individuals with low (<2%, n=313) and high (>2%, n=62)) autozygosity. On the right, the AD *de novo* rate has been used for normalization. Individuals with high autozygosity had a higher relative burden of recessive diseases, mainly due to the presence of homozygous pathogenic variants. Box plots present the median as the center line, the upper and lower quartiles as box limits, and the 1.5-times the interquartile range as whisker length (in the style of Tukey). AD: autosomal dominant inheritance, variant inherited or of unknown origin; AD (*de novo*): autosomal dominant inheritance with *de novo* variant; AR (comp het): autosomal recessive inheritance with compound heterozygous variants; AR (hom): autosomal recessive inheritance with homozygous variant; mt: mitochondrial inheritance; XL: X-linked inheritance.

### Recessive disease burden

The second-largest proportion of solved cases involved an autosomal recessive (AR) mode of inheritance (125 solved cases, 14.5% of all diagnoses; Figure 3a). In total, 94 of the causative variants in the 125 recessive diagnoses in the present cohort would also have been classified as pathogenic if identified in healthy individuals^13^. The diagnostic yield was considerably higher in patients with presumed consanguinity (low autozygosity 31%, n=1,014 vs. high autozygosity 41%, n=144, p=0.01, Fisher’s exact test), and the composition of the modes of inheritance also differed significantly between the high and low autozygosity groups (Figure 3b). The relative contribution of homozygous variants was significantly higher in the high autozygosity group (73%, n=62 diagnoses) than in the low autozygosity group (2%, n=313 diagnoses) (odds ratio (OR) 111.5, p<0.001, Fisher’s exact test). In contrast, the contribution to disease of *de novo* variants was 13% (n=62 diagnoses) in the high autozygosity group compared to 54% (n=313 diagnoses) in the low autozygosity group (OR 0.2, p<0.001, Fisher’s exact test). Since the *de novo* mutation rate is dependent on parental age but not on autozygosity, the disease prevalence that is attributable to *de novo* variants should be comparable in both groups, and can be used for normalization (Figure 3c). For an inbreeding coefficient of >2%, this suggests a recessive disease burden that is seven-fold higher than for those with lower inbreeding coefficients, which is consistent with previous reports^14–16^.

### Dual molecular diagnoses

For 11 individuals, who represented approximately 2% of all solved cases, molecular diagnoses for two distinct or overlapping disease phenotypes were established (Supplemental Table 2). This group showed a tendency for high autozygosity (43%, n=7, vs. 16%, n=361, p=0.09, Fisher’s exact test) and recessive disorders (41%, n=22 diagnoses vs. 24%, n=488 diagnoses, p=0.08, Fisher’s exact test). The detected percentage of dual diagnoses (2%, 11 of 499 solved cases) is consistent with both the enrichment of high autozygosity and recessive disorders in this group, and earlier reports^17,18^.

### Enrichment of ultra-rare diagnoses

For the 499 individuals in whom ES led to a molecular diagnosis, a total of 549 disease causing variants were identified in 362 different disease-associated genes, as well as structural variants affecting 14 genomic regions (Supplemental Table 1). This plethora of diagnoses suggests that each specific genetic disorder had a very low prevalence. To clarify this, the results were compared with the total number of (likely) pathogenic ClinVar submissions for the respective genes (Figure 4a). The first quartile of ClinVar variants corresponds to the more frequently identified rare diseases, and contains 89,707 variants assigned to 42 genes. In the group of 499 individuals with a molecular diagnosis in the present cohort, only 24 patients and 10 different disease-associated genes fell into this first quartile. In contrast, the majority of the present 499 patients (corresponding to 113 different disorders) were assigned to the fourth quartile, which contains disease genes with the least ClinVar submissions (Figure 4b). Notably, almost half of the diagnoses assigned in the present cohort were only established in the last decade (Figure 4c). A comparison to a cohort of comparable size^19^ revealed a significantly different distribution with respect to the years in which the phenotype was first associated with the respective disease-causing gene (Kolmogorov-Smirnoff test, p<0.001, Supplemental Figure 5).

**Figure 4:**
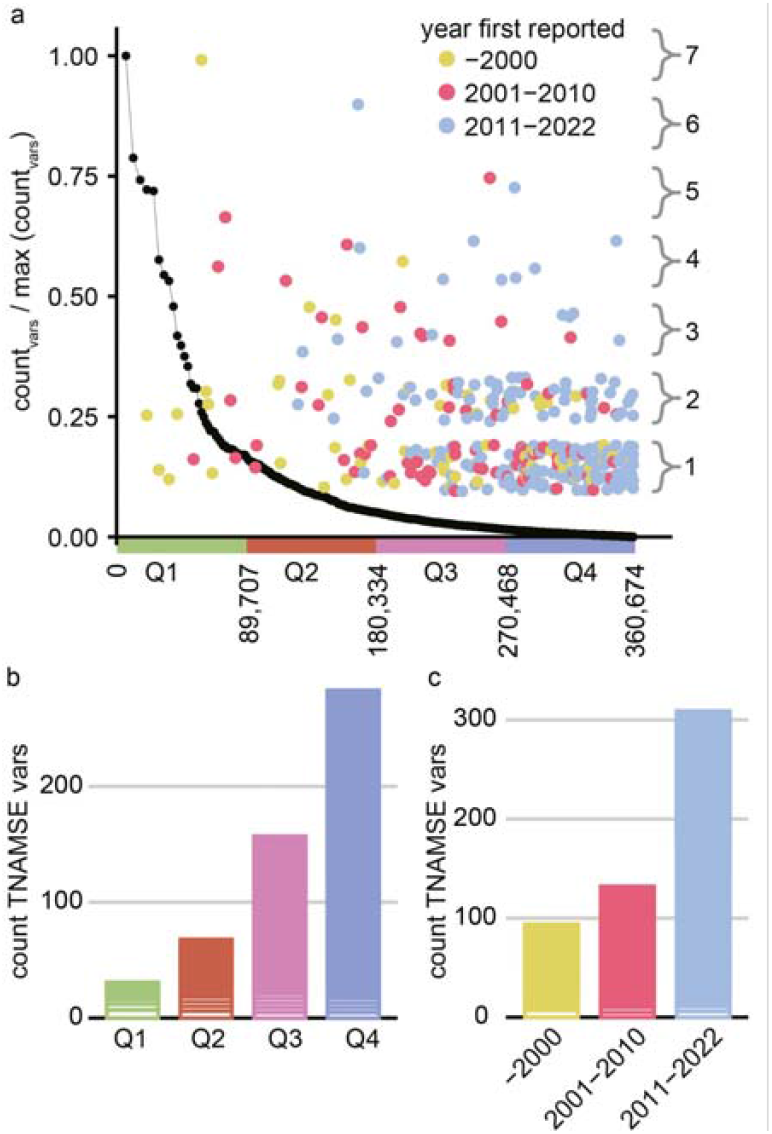
Most variants identified in TRANSLATE NAMSE ES cohort cause ultra-rare disorders that were first associated with a gene in the last decade. a) Comparison of (likely) pathogenic variants per gene, as submitted to ClinVar or identified in TRANSLATE NAMSE. Genes are ordered from left to right according to a decreasing frequency of ClinVar submissions. The black line corresponds to the complementary cumulative distribution (1-CDF; cumulative distribution function) of ClinVar submissions. Diagnostic variants in TRANSLATE NAMSE (counts displayed on the right axis) were plotted as dots above their respective gene and in the color corresponding to the year in which the gene was first described as being associated with the respective disease. (b) Variant counts in TRANSLATE NAMSE in genes with high (first quartile, Q1) to low (Q4) variant counts per gene in ClinVar. The genes in Q1-Q4 each cover approximately 1/4 of the likely or confirmed pathogenic variants in ClinVar, as shown on the x-axis in a). Variants in the same gene are grouped in horizontal blocks. c) Bar graph showing the number of variants relative to the time interval in which the gene was first described as being associated with the respective disease. TNAMSE: TRANSLATE NAMSE; vars: variants.

### Novel diagnostic-grade genes (DGG) and candidates

In 65 cases, most of which concerned a neurodevelopmental phenotype (77%), indications for novel disease associations were identified for a total of 57 genes. Moderate evidence was generated for 23 of these 57 candidates, and high evidence was generated for the remaining 34 (Supplemental Results and Supplemental Table 3). A total of 17 candidate genes with high evidence are currently undergoing further investigation, mostly within the framework of international projects. A total of 17 genes (12 associated with autosomal dominant inheritance, 5 with autosomal recessive inheritance) have subsequently acquired

DGG status through international cooperation^20–32^. In comparison with pathogenic variants in previously known disease-associated genes, the present candidate gene set showed a higher proportion of missense variants. This is probably attributable to the fact that the classification of missense variants is more challenging (Supplemental Table 3).

### Functional assays

For 18 cases that were classified as uncertain or unsolved after initial ES, multi-omic assays were performed, i.e., an analysis of the methylome (n=4), the proteome (n=3), or the transcriptome (n=14). Epigenetic signatures, as derived from methylome analyses, clarified the status of *de novo* missense variants as likely benign in one case, and as pathogenic in three. This is exemplified by a case with a missense variant in *KMT2D* (Supplemental Case Reports)^35,36^. Variants in *MDH2* were reclassified to pathogenic, based on a proteome analysis of patient-derived fibroblasts (Supplemental Case Reports), while results were inconclusive in two unsolved cases. In 13 unsolved cases, RNA sequencing was performed but could not identify transcriptome alterations that lead to the identification of causative variants. Thus, in 5/18 cases, complementary assays facilitated variant reclassification, and highlighted the importance of variant validation strategies in diagnostics for suspected rare genetic diseases (Supplemental Case Reports)^37–39^.

### Predicting the diagnostic yield of ES using machine learning (YieldPred)

Analyses were then conducted to investigate whether the phenotype predicted the diagnostic yield of ES. For this purpose, a least absolute shrinkage and selection operator (LASSO) analysis for binary outcomes was performed. To reduce the phenotypic dimension and to increase interpretability, HPO terms were first aggregated into 49 non-overlapping phenotypic groups. These phenotypic groups were used as predictors in the LASSO analysis. The resulting model was able to discriminate between solved and unsolved cases (Supplemental Figure 3, area under the curve (AUC)=0.67, 95%-confidence interval=0.61-0.74, on a held-out test set of the ES cohort, n=321), and yielded the HPO groups “dysfunction of higher cognitive abilities”, “hematological abnormalities”, and “ataxia” as very influential predictors in terms of the establishment of a molecular diagnosis via ES (Figure 5a). This model was then used to build the machine learning tool YieldPred, which was validated using an external cohort (Supplemental Figure 3, n=753, AUC=0.58). YieldPred has now been made available as a web service that can be used to estimate the diagnostic yield of ES on the basis of the phenotypic features of a given patient (https://translate-namse.de).

**Figure 5:**
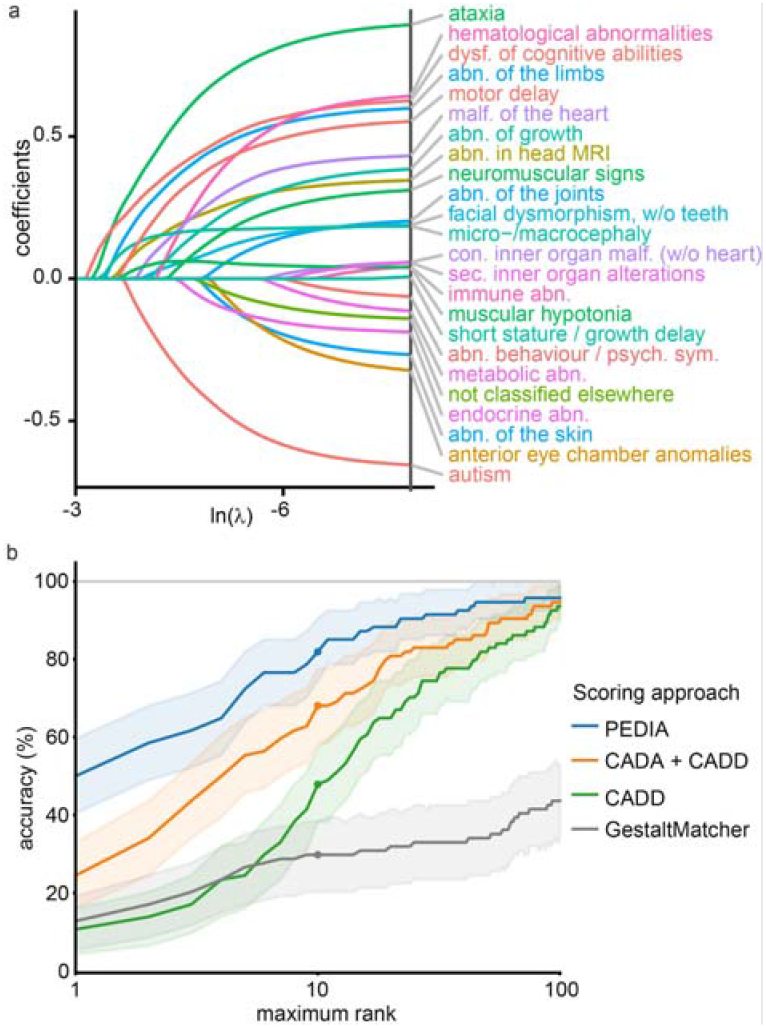
Machine learning identifies features relevant to the diagnostic yield and can support variant prioritization. a) The coefficient paths of regression analysis using the Least Absolute Shrinkage and Selection Operator (LASSO) are shown. The more to the left [lower ln(λ)] a coefficient path starts to deviate from the x-axis, the more informative the corresponding feature is in terms of predicting the diagnostic yield. Features with positive coefficients increase the diagnostic yield. In contrast, features with negative coefficients render a monogenic cause less likely. For example, dysfunction of higher cognitive abilities and ataxia are associated with a higher diagnostic yield (clinical features are colored according to their higher-order HPO groups, for details, see Supplemental Material and Methods). An algorithm to predict the diagnostic yield (YieldPred) was developed based on these data and can be found online (https://translate-namse.de). b) The performances of variant prioritization approaches were compared. All disease-associated genes were ranked using the respective variant prioritization method. Subsequently, the proportion of cases detected with the correct disease-associated gene (sensitivity) was shown as a function of the number of disease-associated genes considered, beginning at the top score. The following four approaches for variant prioritization were tested in solved cases from the PEDIA cohort (n=94): 1) only a molecular pathogenicity score (CADD^86^) with top-10 accuracy of 48%; 2) feature-based score (CADA^85^) in addition to CADD with top-10 accuracy of 68%; 3 and 4) A gestalt score from facial image analysis (GestaltMatcher^40^) alone or in addition to both CADD and CADA referred to as PEDIA score^41^ with top-10 accuracy of 82%. Note that bootstrapped 95% confidence intervals are indicated by the lighter shading around the lines. MRI: magnetic resonance imaging; abn.: abnormality, con.: congenital dysf.: dysfunction; psych.: psychiatric; sym.: symptoms; sec=secondary; CADA: Case Annotation and Disorder Annotation; CADD: Combined Annotation-Dependent Depletion; PEDIA: Prioritization of Exome Data by Image Analysis

### Improving the efficiency of variant interpretation using facial image analysis (PEDIA)

A total of 224 of the 1,577 patients had also provided written informed consent for the evaluation of their facial images with the AI tool GestaltMatcher^40^ and the use of the results (gestalt scores) in exome variant interpretation (PEDIA, prioritization of exome data by image analysis)^41^. In 94 of these PEDIA subcohort cases, a molecular diagnosis was established. For 81 of these 94 cases, the gestalt scores improved prioritization results, that is the correct diagnosis was ranked higher. In general, the PEDIA approach can contribute to prioritization efficiency, provided that: 1) the clinical features of the underlying disorder include facial dysmorphism; and 2) molecularly solved cases are already part of the GestaltMatcher Database^40^ (https://db.gestaltmatcher.org/). In the present PEDIA subcohort, for 81 cases, representing 68 different disorders, one or more previously solved cases were phenotypically so similar that the gestalt score for the associated disease gene resulted in a higher ranking for the pathogenic variant than prioritization approaches that do not make use of image analysis.

Four different variant prioritization approaches involving genotype-based and/or phenotype-based scores were analyzed, and their respective accuracy rates compared. For the PEDIA approach, the correct disease-associated gene was listed among the top ten suggestions in 82% of the cases. The PEDIA approach outperformed prioritization by either a molecular score (CADD^42^) or GestaltMatcher only (Figure 5b). PEDIA was also more efficient than combining a molecular score with a feature score (CADD+CADA), an approach which can be considered routine in ES analysis.

Based on these results and the extension of the TRANSLATE NAMSE study beyond the initial 3-years, the PEDIA workflow was implemented at further sites. The ES data of another 149 patients were then analyzed. In this additional cohort, a molecular diagnosis was established in 69 patients, and a top-10 accuracy of 83% was achieved using the PEDIA score (Supplemental Figure 4).

In some cases, the gestalt scores were particularly suggestive, and facilitated the identification of otherwise challenging pathogenic variants. For instance, in a patient with a very high gestalt score for Koolen de Vries syndrome, a 4.7 kb *de novo* deletion affecting *KANSL1* was detected^43^. Other case reports of particular interest are described in the Supplemental Materials (Supplemental Figure 5).

### Diagnoses with causal therapeutic implications

For five patients in the TRANSLATE-NAMSE cohort with a molecular diagnosis, individualized treatments, or therapies directed against the mechanism of the disease could be initiated^42^. A patient with metachromatic leukodystrophy (MLD) due to pathogenic variants in arylsulfatase alpha (ARSA) was treated with autologous CD34+ cells that were transduced *ex vivo* using a lentiviral vector encoding ARSA^43^. The gene therapeutic approach with atidarsagene autotemcel has been authorized by EMA in the EU since 17 December 2020. A patient with pyruvate dehydrogenase E1-alpha deficiency due to a *de novo* variant in *PDHA1* and another patient with GLUT1-deficiency due to pathogenic variants in *SLC2A1* were treated with a ketogenic diet. In a patient with cerebral creatine deficiency syndrome 1, due to a missense substitution in *SLC6A8*, supplementation with creatine was started. In a patient with congenital disorder of glycosylation of type IIc, due to a homozygous missense variant in *SLC35C1* the fucosylation deficiency was treated by oral fucose supplementation^44^.

## Discussion

Reducing the time to diagnosis from several years to less than one year is highly relevant in terms of both prognosis and the targeted use of healthcare resources, since the number of approved therapies for rare diseases in which early treatment is associated with better outcomes is now increasing^45^. Establishing a molecular diagnosis quickly will require the implementation of frameworks within healthcare systems that are dedicated to patients with rare diseases. The novel diagnostic approach evaluated in TRANSLATE NAMSE was the practical realization of such a concept. The present investigation suggests that a combination of a structured clinical assessment by an MDT and subsequent ES may reduce the diagnostic delay, and that ES was particularly beneficial for patients with ultra-rare genetic disorders. These findings are consistent with reports from other healthcare systems^19,46–50^. In several patients from the present cohort, molecular diagnoses also resulted in a change of clinical management to a causal or even curative approach to therapy as described above. These cases emphasize the fact that molecular genetic diagnoses are essential in terms of the development of personalized treatments or therapies that are directed against the underlying disease mechanism. The systematic, consortium-based collection of molecular and clinical data represents the first necessary milestone towards achieving this goal. Particularly in the case of ultra-rare disorders, the collection of these data requires additional international collaborative efforts.

The present analyses identified a large number of individuals who carried variants that indicated a novel disease-gene association (12% of solved cases), which highlights the fact that the analysis of ES data should not be limited to known disease genes. Establishing novel gene-disease associations and conducting functional analyses for the reclassification of variants of uncertain significance (VUS) are time-consuming and highly complex endeavors^53^. Hence, from the present logistical perspective, such analyses are easier to perform in a research context than within the routine diagnostic context of clinical practice. However, these findings are of crucial importance for affected individuals and their families. Thus, from a teleological perspective, in some rare disease cases, boundaries separating diagnostics and research are somewhat blurred. Therefore, in the tertiary, academic setting, collaboration between experts from diagnostics and research is highly relevant for patients with suspected ultra-rare diseases and a lack of definitive diagnostic findings.

The present study demonstrated that ES and MDT improve diagnostics for patients with rare diseases within the German national healthcare system. On the basis of the present data, in 2021, ES was included in the list of standard medical services offered to patients with suspected rare diseases who were referred to German CRDs. For all the patients that are still awaiting a molecular diagnosis new multi-omics approaches are promising, but they also costly. Therefore, in a complex healthcare system, these tests compete with other analyses, and their efficiency and efficacy in establishing a diagnosis should be evaluated in the future.

Another aim of the present study was to determine whether complementary AI and machine learning approaches would facilitate diagnostic effectiveness and efficiency in the ES cohort. The PEDIA analyses showed that AI–powered next-generation phenotyping increased the efficiency of ES data analysis. However, not every case in the present cohort was solved via ES. Therefore, to predict the diagnostic yield of ES on the basis of patient phenotype, the machine learning model YieldPred was developed. YieldPred can be used to estimate the likely efficacy of ES, i.e., the probability that ES will lead to the establishment of a molecular diagnosis in a given patient with a suspected rare disease. Users can specify the age, sex, and assigned HPO terms of their patient. Low scores in YieldPred despite a high likelihood of a monogenic cause, e.g. a strong family history, could justify the selection of an even more comprehensive test, such as long-read genome sequencing.

Besides the ability to select the appropriate genetic test for diagnosing a disease, a core competence of a clinical geneticist is to estimate disease risk in the offspring of healthy individuals. In addition to the relatedness of the partners, the burden of heterozygous pathogenic variants in recessive genes - which can vary considerably depending on demographics^54–57^ - could play an increasingly important role in family planning. In a total of 94 of the 125 cases with recessive molecular diagnoses, the causal variants would also have been classified as (likely) pathogenic if they had been identified in healthy individuals^13^. This also means that if the parents of pediatric patients with a recessive disorder in the present cohort had undergone ES to determine their carrier status, three out of four of these couples could have received appropriate genetic counseling concerning disease risk in future offspring, which supports the argument for extended screening^58^.

Two notable findings of the present analyses were that in comparison to ClinVar and a previously reported rare disease cohort of similar size^19^, the TRANSLATE NAMSE cohort was significantly enriched for ultra-rare disorders (Figure 4a, Supplemental Figure 6a, b), and that a large number of recently described gene-disease associations were found (Figure 4c, Supplemental Figure 6c)^1,8,19,59^. In our opinion, this accumulation of ultra-rare diagnoses is not explained solely by prior negative genetic testing. Rather, it reflects the value of the MDTs in terms of guiding the diagnostic process. Furthermore, the fact that a large number of the established diagnoses have only become possible in recent years as a result of increasing medical genetic knowledge (Figure 4c) highlights the importance of data reanalysis^60,61^.

It would be desirable for all individuals with a suspected monogenic disorder for whom no definitive diagnosis can yet be established to have the option of participating in large-scale genomic diagnostic and research initiatives. We present TRANSLATE NAMSE as the German framework that organizes diagnostics for patients with ultra-rare diseases with a backbone of case conferences in MDTs in academic centers for rare diseases. TRANSLATE NAMSE represents the first national-level project for undiagnosed patients in Germany, and the future expansion of the network on both the national and international level is planned.

In summary, the results of the present study demonstrate that our novel, structured diagnostic concept facilitates the identification of ultra-rare disorders on a national level, provides undiagnosed patients with the opportunity to participate in international research, and represents a platform for data sharing that facilitates the development of machine learning and AI tools to improve diagnostic effectiveness and efficiency.

## Online Methods

### Study design

#### Enrollment, research ethics, and consent

A detailed description of the TRANSLATE NAMSE project is provided elsewhere.^9,62^ In brief, participants for TRANSLATE NAMSE were recruited between January 2018 and December 2020 from a total of 10 German CRDs (Berlin, Bochum, Bonn, Dresden, Duisburg/Essen, Hamburg, Heidelberg, Kiel/Lübeck, Munich, and Tuebingen). Overall coordination of the recruitment process was performed by the Institute of Public Health Berlin. The study was approved by the internal review board of each participating institution. All patients or their legal guardians provided written informed consent prior to inclusion. The inclusion criteria for TRANSLATE NAMSE were the lack of a definitive diagnosis and the clinical suspicion of a rare disease. The medical records and family history of each individual were evaluated by a multidisciplinary team (MDT), which comprised at least board-certified physicians of two specialities with domain-specific expertise. For each individual, the respective MDT then made recommendations concerning diagnostics and further clinical management. To make the recommendation of ES a board-certified human geneticist was additionally required within the MDT. For example, strong criteria for the indication of ES were congenital malformations, a syndromic phenotype, a positive family history suggestive of a monogenic disease as well as lack of absence of an alternative test with a comparable suspected diagnostic yield. A total of 1,577 patients (268 adult and 1,309 pediatric) from the TRANSLATE NAMSE cohort were referred for ES on the recommendation of the MDT at the respective CRD (ES cohort). The phenotypic and molecular genetic data of these 1,577 patients were evaluated in the present analyses.

### Clinical and laboratory phenotype data

Clinical and laboratory phenotype data were transferred to the sequencing laboratory in the form of hard-copy case report forms or as online data capture applications (Face2Gene Clinic). Online data capture allowed the free entry of Human Phenotype Ontology (HPO) terms. Data from hard-copy report forms and free-text entries were transformed into HPO terms. The phenotypes reported in the present study are those that were reported to the sequencing laboratories. On the basis of the leading presenting clinical feature, each case was assigned to one of six major disease groups (Supplemental Figure 1b). This allowed a more definitive statement on diagnostic yield in relation to the clinical features of the patient. In the subsequent analyses, all assigned HPO terms (n=1,649) were compiled and divided into higher-order groups (n = 12) and subcategories (n = 49) by expert clinicians. Therefore, patients were additionally assigned to at least one higher-order group as well as at least one subgroup. To assign a patient to an HPO-defined group, the patient had to have at least one of the HPO terms belonging to the respective group. The following higher-order groups were defined: 1-neurodevelopmental, 2-neuromuscular, 3-seizures, 4-growth disorders, 5-facial dysmorphism, 6-abnormality of connective tissue, 7-congenital malformations, 8-endocrine and metabolic abnormalities, 9-immune and hematological abnormalities, 10-sensory organ alterations, 11-abnormal findings on brain magnetic resonance imaging, 12-others. Within the respective higher-order groups, HPO-terms were further assigned to subcategories (n=49) (https://github.com/Ax-Sch/TNAMSE_geno_pheno/blob/main/resources/hpo_categorization_19_12_2022.tsv).

### DNA sequencing

EDTA-treated whole-blood samples or saliva kits were delivered to one of the five participating sequencing centers (Berlin, Bonn, LMU Munich, Munich, Tuebingen) for further processing: After DNA extraction, fragment size and purity were assessed. If the DNA fulfilled all quality criteria, the sample was submitted for sequencing. ES was performed on exon targets that were isolated using capture and either Agilent SureSelect Human All Exon kits v6 or v7 (Agilent Technologies, Santa Clara, USA), or the Human Core Exome kit (Twist Bioscience, San Francisco, USA). One microgram of DNA was sheared into 350-to 400-bp fragments, which were then repaired, ligated to adaptors, and purified for subsequent polymerase chain reaction (PCR) amplification. Amplified products were then captured by biotinylated RNA library baits in solution, in accordance with the manufacturer’s instructions. Bound DNA was isolated with streptavidin-coated beads and reamplified. The final isolated products were sequenced using the Illumina HiSeq 2500 or NovaSeq 6000 sequencing system and 2x100-bp paired-end reads (Illumina, San Diego, CA). All five sequencing centers ensured a coverage of over 20x in over 95% of the RefSeq target region.

### ES data-processing pipeline

At each of the five sequencing centers, ES processing pipelines were established according to best practice guidelines. The DNA sequence was mapped to the published human genome build GRCh37 reference sequence using Burrows-Wheeler Aligner (BWA). The most up-to-date version at the time of sequencing was used, progressing from BWA v0.7.11 through to BWA-Mem v0.7.17^63,64^. Single nucleotide variants (SNVs) and small insertions and deletions (indels) were detected with HaplotypeCaller and SAMtools v.0.1.7^65,66^. Mitochondrial DNA variants were assessed using data from ES^67^. Copy number variations were detected using ExomeDepth and Pindel on short-read data, or prior to ES by array CGH^68,69^.

Variants were annotated using VEP^70^ or Jannovar^71^ and analyzed in VarFish^72^ or EVAdb (https://github.com/mri-ihg/EVAdb), depending on the center.

The population background of each individual was estimated with peddy^73^ . This revealed that the cohort was of predominantly European origin (Supplemental Table 1, Supplemental Figure 7).

Autozygosity was estimated using RohHunter, bcftools/roh, or a sliding-window framework^74–76^. A small subset of samples was run on all three tools, and this yielded comparable results for autozygosity. A threshold of 2% was used to assign patients to a high or a low autozygosity group^14^ (Supplemental Figure 7).

The variants identified in ES were assessed in accordance with the standards and guidelines of the ACMG for the interpretation of sequence variants^77^. At least two physicians or experts in molecular genetics participated in the assessment of the variants. Finally, all variants that were potentially disease-causing (pathogenicity class 3-5), and actionable secondary findings were reported to the respective patients.

Cases in which no diagnosis could be established in a known disease-associated gene were included in national and international studies for the discovery of novel disease etiologies e.g., via the MatchMaker Exchange (MME) Network.^78,79^ Variants with a high likelihood of being disease-causing, e.g., those with loss of function or high pathogenicity scores, or those that had arisen *de novo*, were shared through MME in order to identify similar patients.^80,81^

### Statistical Analyses

All statistical analyses were conducted in R (version 4.1.2).^82^ Proportions were tested using a two-sided Fisher’s exact test. The significance level was set to α = 0.05 and p-values were corrected via Bonferroni correction if necessary.

### Visualization of phenotype space using uniform manifold approximation and projection (UMAP)

First, data on known diseases and their clinical features were downloaded from the HPO website (https://hpo.jax.org/app/download/annotation, file: genes_to_phenotype.txt, downloaded on 10 April 2021). The disease data were merged with the data of the 1,577 individuals from TRANSLATE NAMSE by treating each disease-ID as one individual. Similarities in HPO terms between all pairs of individuals were then calculated using the R package ontologySimilarity (version 2.5). The similarities were then converted to a distance matrix and projected into a four-dimensional space using UMAP. Subsequently, the first two dimensions of this projection were plotted using ggplot2 (version 3.3.4).

### Variants amenable to carrier screening

In cases with autosomal recessive inheritance, disease-causing variants in ClinVar were queried in January 2017 (beginning of the project) in order to take into account the state of knowledge available at the time of analysis. Variants were classified as amenable to carrier screening if they were classified as pathogenic or likely pathogenic in ClinVar, or if they were predicted loss of function variants that were not predicted to escape nonsense-mediated mRNA decay. In compound-heterozygous inheritance, both variants were required to be (likely) pathogenic.

### Comparison of disease-associated genes reported in TRANSLATE NAMSE to those reported in other cohorts

In the German healthcare system, genetic testing of the more frequent rare disorders, e.g., retinitis pigmentosa, or hearing impairment, is performed using gene panels.

For a comparison with Turro, *et al*., all disease-associated genes were first ranked according to the frequency of submissions of pathogenic and likely pathogenic variants to ClinVar. Disorders caused by genes in the first quartile of the ClinVar gene distribution, such as *USH2A, ABCA4*, and *BMPR2*, are more prevalent than phenotypes associated with genes in the fourth quartile. In addition, the year in which phenotype-gene associations had first been reported was determined in order to assess when a diagnosis could first have been established. The characteristics of the variants identified in the TRANSLATE NAMSE ES cohort were then compared to those identified in a cohort reported by Turro, et al. in 2020.

Turro, *et al*. subjected 9,802 individuals with a suspected rare disease to genome sequencing and reported pathogenic or likely pathogenic variants in 1,138 cases.^19^ Around a quarter of these variants were assigned to genes with a high disease prevalence (Supplemental Figure 8). In contrast, most disease-associated genes identified in the TRANSLATE NAMSE cohort were ultra-rare, and more frequent diagnoses were underrepresented.

### Novel disease candidate genes

Sequence data from the unsolved cases were analyzed for variants in potential novel disease candidate genes. The following mandatory criteria for novel disease candidate genes were defined: (1) the gene had shown no previous robust association with any human phenotype; (2) no other clearly causative disease explanation was found; (3) the allele frequency of the respective variant was below the minor allele frequency (MAF) cutoff or the variant was absent in controls; (4) inheritance was in accordance with the phenotype in the family and/or the variant co-segregated with the disease in multiple affected family members. As in the ClinGen approach and as suggested by others, characteristics, including gnomAD constraint metrics, inheritance, and functional data, by which the level of evidence for the manually identified candidate genes could be assessed were defined^34,53,83^ (Supplemental Table 3). An evidence score was then calculated, which could reach a maximum value of 8. Three of the nine criteria can only be applied to genes with an autosomal dominant mode of inheritance (*de novo* status and gnomAD constraint metrics), rendering the score less informative for autosomal recessive inheritance. For autosomal dominant inheritance, a score of 1-3 was ranked as medium evidence, and a score of 4 and above as high evidence. For recessive inheritance, a score of 3 or above was ranked as high evidence, and a score of below 3 was ranked as medium evidence. Genes first published as disease-associated during the course of TRANSLATE NAMSE were classified as novel DGG.

### Diagnostic yield prediction (YieldPred)

The ES cohort (n=1,577) was randomly divided into a training set comprising 1,256 cases (399 solved, 32%), and a test set comprising 321 cases (99 solved, 31%). The binary status of a case (1=solved, 0=unsolved) was regressed on the 49 HPO-defined subcategories (cf. clinical and laboratory phenotype data) using LASSO for binary outcomes with the logit function as a link function (R package glmnet, version 4.1-4), and by controlling for age (adult/child), sex (male/female), sequencing laboratory, and the use of the PEDIA workflow. Variable selection was applied on the 49 HPO-defined subcategories only. The model was fitted on the training set, and the penalty parameter was tuned via ten-fold cross-validation. The resulting model was then applied to the test set, and its predictive performance was evaluated using the Receiver Operator Characteristics (ROC) curve. The model was then refitted on the whole ES cohort of 1,577 cases and the resulting model was validated on an external and independent cohort (n=753, 545 solved, 72%, Supplemental Table 4). This validation cohort was recruited by the Technical University of Munich and all individuals consented in the scientific use of their phenotype and genotype data. The final tool YieldPred was provided as a web service, where users can specify the age, sex, and assigned HPO terms of their patient, while the remaining confounders are estimated via the mean confounder values of the TRANSLATE NAMSE ES cohort.

### PEDIA analysis

Prioritization of exome data by image analysis (PEDIA) integrated the facial image and clinical feature analysis with exome data analysis^41^. For each patient, a frontal facial image, clinical features encoded in HPO terminology, and exome sequencing data were available for analysis.

The PEDIA approach was used, in which the facial image analysis was analysed by GestaltMatcher^40^. GestaltMatcher was trained on 6,354 frontal images with 204 different disorders in order to learn the respective facial dysmorphic features, and it further encoded each image into a 512-dimensional facial phenotype descriptor. The model ensembles and test-time augmentation were later used to generate 12 512-dimensional facial phenotype descriptors for each image^84^. The similarity between two patients can be quantified by averaging 12 cosine distances of the facial phenotype descriptors. For each test image, a list of similarity scores for 816 disease-causing genes were obtained. To convert HPO terms of individual patients into feature scores for each gene, the CADA approach was used^85^. For the exome data, each variant was annotated with a version 1.6 CADD score^42^. After filtering out the common variants, the highest CADD score for each gene was taken.

In this analysis, benchmarking was performed on two cohorts: PEDIA-subcohort and validation-cohort. The PEDIA-subcohort consisted of a subset of 224 of the 1,577 ES patients (194 pediatric, 30 adult). Of these, 94 had a molecular genetic diagnosis (86 pediatric, 8 adult). After the end of the three year TRANSLATE NAMSE recruitment period, a further 149 patients were enrolled and used as a validation cohort. In the validation cohort, 69 out of 149 patients were solved cases. All facial images analyzed in the present study can be accessed in GestaltMatcher Database (https://db.gestaltmatcher.org/) by the GMDB ID in Supplemental Table 1 and Supplemental Table 5. For each patient, each gene had a GestaltMatcher score, a CADA score, and a CADD score. These three scores were the input of the PEDIA approach. The output for each patient was a list of genes, and each gene had a PEDIA score. The genes were then prioritized by ranking the PEDIA scores in descending order. To benchmark the performance, top-*k* accuracy was used, as calculated by the percentage of the patients with the disease-causing gene ranked in the top-*k* position. Finally, the top-1 to top-100 accuracies of the two cohorts (PEDIA-subcohort of the ES cohort and validation cohort) were reported.

## Supporting information

Rebuttal

## Data Availability

All data produced in the present work are contained in the manuscript and online at https://translate-namse.de

## Data availability

The corresponding author will comply with all requests for materials that are not presented in the extended data and Supplemental information, after verification of whether the request is subject to any patient confidentiality obligation. Patient-related data not included in the article may be subject to patient confidentiality.

The genotype and phenotype data of 5,652 participants from the TRANSLATE NAMSE project can be accessed by following the procedure outlined at https://www.translate-namse.de Reported alleles and their clinical interpretation have been deposited in ClinVar using the following submitters:

Institute for Genomic Statistics and Bioinformatics (University Hospital Bonn):

https://www.ncbi.nlm.nih.gov/clinvar/submitters/507028/, https://www.ncbi.nlm.nih.gov/clinvar/submitters/508040/

Institute of Human Genetics, Klinikum rechts der Isar (Technical University Munich): https://www.ncbi.nlm.nih.gov/clinvar/submitters/500240/,

Institute for Medical Genetics and Human Genetics (Charité-Universitätsmedizin Berlin): https://www.ncbi.nlm.nih.gov/clinvar/submitters/505735/,

Institute of Medical Genetics and Applied Genomics (University Hospital Tübingen): https://www.ncbi.nlm.nih.gov/clinvar/submitters/506385/.

Genomics Facility (Ludwig-Maximilians-Universität München): https://www.ncbi.nlm.nih.gov/clinvar/submitters/507363/

## Code availability

The study’s landing page (https://www.translate-namse.de) redirects to a web service for the prediction of the diagnostic yield and the code repository. All source codes are available under a creative commons license.

## Acknowledgements

We thank all patients and families for their cooperation. We thank Christine Schmael for proof-reading of the manuscript. Magdalena Danyel, Henrike Sczakiel and Martin Attah Mensah are participants in the BIH-Charité (Digital/Junior) Clinician Scientist Program, which is funded by Charité – Universitätsmedizin Berlin and the Berlin Institute of Health (BIH). Felix Boschann is a participant in the Clinician Scientist Program (CS4RARE) funded by the Alliance4Rare and associated to the BIH Charité Clinician Scientist Program.

## Notes

### Competing Interest Statement

The authors have declared no competing interest.

### Funding Statement

This study was funded by Innovationsfonds https://innovationsfonds.g-ba.de

### Author Declarations

The institutional review board of University Hospital Bonn, Germany approved the study (312/17)

