## Supplemental Material for "Next-generation phenotyping integrated in a national framework for patients with ultra-rare disorders improves genetic diagnostics and yields new molecular findings"

**Contents**

Supplemental Results	1

Novel disease candidate genes	1

Gene length calculation	2

Parental Mosaicism	3

Secondary Findings	4

Case reports of particular interest	4

Supplemental Figures	8

Supplemental Tables	15

References	17

### Supplemental Results

#### Novel disease candidate genes

A total of 57 different candidate genes in 65 affected individuals from 63 unrelated families had an evidence score of ≥1 (Supplemental Data Table 3). The inheritance pattern was autosomal dominant for 41 genes (evidence score range 1-7); autosomal recessive (AR) for 15 genes (evidence score range 1-5); and X-chromosomal for one gene (evidence score 1). Sixteen genes have subsequently acquired diagnostic-grade gene (DGG) status. Selected cases are described below, most of which have been described in detail in previous publications.

*SMARCA5*

In *SMARCA5*, a gene that encodes a chromatin remodeler, we identified *de novo* variants in two patients with neurodevelopmental delay and similar dysmorphic features. The phenotype-gene association was strengthened by the identification of 10 additional cases from other cohorts. Rescue experiments with wildtype transcripts in Drosophila suggested that the identified *de novo* variants had a hypomorphic effect^1^.

*KCNN2*

In another individual with learning disabilities, autism, dystonia, and intention tremor, we identified a *de novo* missense variant in *KCNN2.* This gene encodes a small-conductance calcium-activated potassium channel protein. Interestingly, a preexisting rat model with a missense substitution identical to that found in the affected individual partially mirrors this phenotype, with abnormal locomotor activity and tremors. The identification of nine additional individuals from other cohorts, and the results of functional analyses of the variants with respect to channel function, established *KCNN2* as a dominant disease-associated gene for a neurodevelopmental movement disorder^2^.

*MAPKAPK5*

We delineated a recognizable syndrome with multi-organ manifestations for biallelic truncating variants in *MAPKAPK5* (MAPK-activated protein kinase 5). Patients presented with severe developmental delay, variable brain anomalies, congenital heart defects, facial dysmorphism, and a distinct type of synpolydactyly involving the presence of an additional hypoplastic digit between the fourth and fifth digits of the hands and/or feet^3^.

*OAS1*

We established the gene *OAS1*, which encodes a type I interferon–induced, intracellular double-stranded RNA (dsRNA) sensor that is required for antiviral defense, as a DGG. A *de novo* gain-of-function (GOF) variant that caused dsRNA-independent OAS1 activity was identified in a patient with an autoinflammatory immunodeficiency. The latter was characterized by B-cell and monocyte apoptosis-related hypogammaglobulinemia and pulmonary alveolar proteinosis. Via an international network of experts on inborn errors of immunity, we identified three additional *de novo OAS1* GOF variants in a total of six patients, and demonstrated that allogeneic hematopoietic cell transplantation can be applied as a curative approach.

#### Gene length calculation

Coding sequence length was calculated on the basis of Consensus Coding Sequences (CCDS; CCDS.current.txt file downloaded from https://ftp.ncbi.nih.gov/pub/CCDS/current_human/ on 22 July 2021). A list of genes that cause Mendelian disease according to Online Mendelian Inheritance in Man database (OMIM) (n=4,369) was obtained using the files genemap2.txt and mimTitles.txt, as downloaded on 15 July 2021. Supplemental Figure 9 displays boxplots comparing: all genes (n=18,598); OMIM disease-causing genes (n=4,369); TRANSLATE NAMSE ES cohort disease-associated genes (n=330); and TRANSLATE NAMSE ES cohort research genes (n=24). For known disease-associated genes in the TRANSLATE NAMSE ES cohort (TN_Diagnostic), and genes for which a novel disease association was found (TN_Research), the mean coding length was significantly longer than for all genes or for all OMIM genes.

#### Parental Mosaicism

Amongst the 228 solved cases that were attributable to *de novo* variants, three probable or certain parental mosaics affecting at least the germ cells were detected. This was achieved via visual inspection of all parental sequence reads in Integrative Genomics Viewer (IGV) for the presence of the corresponding variant reads (≥ 1). In two of the three families, the unequivocal presence of low-level parental mosaicism was not confirmed, since in family 2, this was only hinted at by a single parental read, while in family 3 no parental mosaicism was demonstrated. However, parental mosaicism is the most probable explanation for recurrence of the “de novo” variant in a second child.

In family 1, which had only one affected member (case 432), a pathogenic, heterozygous nonsense variant in *ASXL3* was detected, leading to the assignment of a diagnosis of autosomal dominant Bainbridge-Ropers-Syndrome (OMIM #615485). The variant was also detected in two out of 69 ES reads from the mother and one out of 60 reads from the father. For the paternal sequence reads, low-quality scores pointed to an artifact. In addition, three unrelated individuals from the same sequencing run carried the variant on one read respectively, suggesting batch contamination. Sanger sequencing revealed the low intensity presence of the variant in maternal lymphocyte DNA, while no evidence for the variant was detected in paternal DNA. Since the latter did not rule out the presence of low-level paternal mosaicism, both parents were sequenced again at a higher coverage. Here, the presence of the variant, and at high quality, was found in 10 out of 177 maternal reads, while no such variant was detected in 197 paternal reads.

In family 2, which included two siblings with intellectual disability (case 435 and his brother), a heterozygous, likely pathogenic variant in *FOXG1* (*FOXG1* syndrome, OMIM #613454) was detected in both individuals. DNA from maternal blood was subjected to ES, and the variant was found in one of 132 reads. No evidence of contamination was found. Sanger sequencing of DNA from both a second maternal blood sample and a maternal buccal swab was then performed. The variant was not detected in either sample.

In family 3, parental mosaicism was established on the basis of the presence of a heterozygous, likely pathogenic variant in the gene *PUF60,* which explained the observed developmental delay in two brothers (cases 443 and 444; Verheij syndrome, OMIM #615583). DNA from the blood of both parents was then subjected to ES. The variant was not detected in either sample, despite a high coverage of 226 and 201 sequencing reads, respectively.

These examples illustrate the frequency of low-level parental mosaicism and its relevance in terms of counseling for recurrence risk. Notably, only two of these three parental mosaics would have been detected in parental next generation sequencing (NGS) reads if the criteria proposed by Gambin and colleagues had been applied, according to which only variants that are present in at least two or more reads and that are absent in reference databases are classified as candidate low-level mosaic single nucleotide variants^4^. The findings in family 3 again demonstrate that the apparent absence of a variant - even in high-coverage parental NGS data – does not exclude gonadal mosaicism.

The detection in the ES analysis of three probable or proven parental mosaics among 228 patients would correspond to a frequency of 1.3%. This is in agreement with two previous studies, which investigated the frequency of low-level parental mosaicism for pathogenic or likely pathogenic variants identified in affected children. Wright and colleagues analyzed 4,293 ES trios and identified and validated eight low-level parental mosaics, six of which were considered likely pathogenic or pathogenic (corresponding to xxx% of the trios)^5^. Cao and colleagues analyzed a cohort of approximately 12,000 samples submitted for clinical ES and identified 0.3% parental mosaics, via analysis of parental NGS reads or via Sanger sequencing of parental DNA^6^.

Our findings highlight the importance of the visual inspection of parental NGS data for low-level mosaicism. In addition, the analyses emphasize the importance of recognizing pedigree constellations that are suggestive of parental gonadal mosaicism.

#### Secondary Findings

Pathogenic (class 5) and likely pathogenic (class 4) variants were only reported to the respective patient if the individual or their legal guardian had previously consented to being informed about secondary findings (SF). The list of actionable genes was based on the recommendations of the American College of Medical Genetics (ACMG, v2.0)^7,8^. However, variants in seven additional genes were reported to patients following discussions within the respective multidisciplinary teams, since it was concluded that these variants fulfilled the criteria for actionable findings (see below). A list of all SFs identified in the TRANSLATE NAMSE ES cohort is provided in Supplemental Table 6.

Medically actionable SFs that were found in genes unrelated to the phenotype, but which had immediate implications for management, were identified in a total of 17 patients (1.1%). In eight cases, the SFs concerned genes included in the ACMG list v2.0^8^. Risk management counseling was provided to the affected individuals, in accordance with the NCCN Guidelines^9^. For the remaining cases, six had variants in the cancer predisposition genes *DICER1, PALB2,* *CHEK2, BRIP1, SBDS,* and *BAP1,* respectively. In view of current recommendations that mutation carriers should avoid exposure to specific agents^10^, two individuals were informed of the SF of X-linked glucose-6-phosphate dehydrogenase deficiency (G6PD; OMIM #300908). Similarly, the reporting of the SF of a pathogenic variant in both *KCNJ11* and in *GHRHR* was of direct clinical benefit to the respective patients and their families, due to the availability of clinical enzyme therapy for maturity onset diabetes of the young type 13 (MODY13, OMIM #616329) and isolated growth hormone deficiency type 4 (OMIM #618157) respectively. Among the ACMG listed genes, the only recurrent findings concerned variants in *BRCA2* and *LDLR,* which were detected in three patients respectively.

In the present multi-center study, the identification and reporting of SF was non-standardized due to cross-center differences in procedures and policies. In particular, cross-center differences were evident with regards to trio ES, when variants were identified that predisposed to a disorder with an onset in adulthood. Here, some centers returned medically actionable results to the adult proband only, and did not automatically inform the respective parents. Centers also differed in terms of both their lists of actionable genes and if they reported heterozygous variants in genes with autosomal recessive inheritance patterns.For some conditions, variants of uncertain significance (VUS) were also reported if a simple clinical test was available for further clarification. One such case involved the identification of a VUS in a gene associated with long QT syndrome, which prompted a recommendation for an electrocardiogram. The number of patients with pathogenic variants in genes from the ACMG list was lower than in previous ES and genome sequencing (GS) studies, which found SFs in 1.2% to 11% of cases^11–15^. This high cross-study variability in SF rates is attributable to: 1) the high discrepancy in terms of the genetic diagnoses that were considered medically actionable; 2) the informed consent procedure (opt-out for non-ACMG SF versus non-opt-out; 3) the applied classification strategy; and 4) the filtering procedures used for variants during exome analysis. This underlines the need for an updated and curated set of actionable genes, as well as updated standards for analysis procedures, the determination of the pathogenicity of variants, and guidelines for returning results to the patient.

#### Case reports of particular interest

*KANSL1* (next-generation phenotyping)

In a female individual (case 393), who first presented at the age of eight years with cognitive impairment, generalized muscle weakness, nevi, iron deficiency anemia, poor fine motor coordination, recurrent respiratory infections, and dysmorphic facies, facial image analysis suggested a high syndromic similarity to Koolen-De-Vries syndrome (Supplemental Figure 5). After inconclusive ES, targeted analyses of *KANSL1* and GS were performed, which revealed a 4.7 kb *de novo* deletion in *KANSL1,* NM_001193466:c.1849-4661_1895del. This example illustrates the value of high gestalt scores, which can be used as an indicator of the need for a targeted search for non-coding disease-causing mutations in the specific genomic region that is associated with the phenotype.^16^

*BICD2* and *TANGO2* (dual diagnosis)

A 13-year-old boy (case 144) with non-consanguineous parents presented with global developmental delay, muscular hypotonia, bilateral optic atrophy, cardiac arrhythmia, hypothyroidism, chronic urticaria, and two episodes of rhabdomyolysis. After an uneventful pregnancy, full term delivery, and normal postnatal development, he had experienced his first atonic seizure at the age of one year. Following bilateral mastoiditis and subsequent mastoidectomy and paracentesis, a first episode of rhabdomyolysis occurred at the age of 16 months, and this later recurred in the context of febrile infections. His development was delayed, with the onset of unassisted walking and first words at the age of 5 years. At the age of 11 years, he was able to speak around 30 words and use 2-word sentences. At 18 months, he experienced an unexplained loss of consciousness in association with gastroenteritis. At 25 months, he developed an intermittent loss of muscle tone, with a fixed gaze, ptosis, and hypersalivation consistent with atonic seizures. At the age of 13 years, the patient had another episode of rhabdomyolysis, this time with cardiac arrhythmia. Since the results of NGS panel sequencing and standard cytogenetic analysis were unremarkable, trio ES was performed. A heterozygous *de novo* missense variant in *BICD2* was detected: *BICD2* (NM_001003800.1): c.2383C>T; p.(Arg795Trp). The disorder *BICD2*-associated spinal muscular atrophy, lower extremity-predominant, 2A (SMALED2A, OMIM #615290) is characterized by motor developmental delay, muscle weakness, muscle atrophy, and distal hyporeflexia, and has an onset in early infancy. A homozygous deletion in *TANGO2* (NM_152906.5):c.57-1743_*10769del, which is associated with AR recurrent metabolic crises with rhabdomyolysis, cardiac arrhythmias, and neurodegeneration (MECRCN, OMIM #616878), was also identified.

*TRDN* and *GYG1* (dual diagnosis)

An 8-year-old boy (case 312) with second-degree consanguineous parents was diagnosed with motor developmental delay, suspected cardiomyopathy, and congenital myopathy after resuscitation for ventricular fibrillation was required at the age of 4 years and 3 months. Following Cesarean section in the second stage of labor at 39+2 weeks gestation, the postnatal period was unremarkable. Motor milestones were delayed. He started to turn at the age of 10 months, sit freely and crawl at the age of 16 months, and walk freely at 26 months. Muscular hypotonia was first documented at the age of 6 months. Cognitive and speech development were unremarkable. Since the results of NGS panel sequencing and standard cytogenetic analysis were negative trio ES was performed. The following homozygous variants were detected: *TRDN* (NM_006073.3):c.508G>T;p.Gly170* and *GYG1* (NM_001184720.1):c.487del; p.Asp163Thrfs*5. Both variants had been described previously ^17^ and were classified as pathogenic. Biallelic pathogenic variants in *TRDN* are associated with AR cardiac arrhythmia syndrome with or without skeletal muscle weakness (OMIM #615441). Biallelic alterations in *GYG1* are associated with two phenotypically overlapping syndromes: glycogen storage disease 15 (GSDXV, OMIM #613507); and polyglucosan body myopathy type 2 (OMIM #616199). To date, reports in the literature indicate a high degree of clinical variability and no specific genotype-phenotype correlation.

*KMT2E* and *DDX3X* (locus heterogeneity in two siblings with NDD)

Two siblings (a 15-year-old girl and a 7-year-old boy) whose parents were first cousins presented with a similar phenotype, comprising global developmental delay, muscular hypotonia, and microcephaly. Duo ES was performed, and the generated data were analyzed under the assumption of an AR disorder, given the parental consanguinity and phenotypic overlap. Duo ES did not lead to the prioritization of any variants that were shared by both siblings. Therefore, ES of the parents with combined data analysis was initiated. While a heterozygous *de novo* stop-loss variant in *KMT2E* (NM_018682.3:c.4743_4744del, p.Phe1582Tyrfs*286) was identified in the brother, a heterozygous *de novo* nonsense variant in *DDX3X* (NM_001356.4:c.841C>T, p.Gln281*) was found in the sister, leading to the assignment of two different diagnoses ("O'Donnell-Luria-Rodan syndrome" and “Intellectual developmental disorder, X-linked, syndrome, Snijders Blok type”, respectively). Reverse phenotyping revealed distinct phenotypes in the two children, each of which aligned with the respective diagnosis. This case description is an excellent example of how parental consanguinity poses a pitfall in variant prioritization, particularly in families with more than one affected child. Moreover, it highlights the need for accurate pre-exome phenotyping, as well as the advantage of a trio approach for the detection of *de novo* variants.

*KMT2D* (functional assays: methylation analysis)

*De novo* heterozygous missense variants were identified in two individuals, and these were initially classified as “likely pathogenic” on the basis of ACMG guidelines^18^: The first case was a 3-month-old, otherwise healthy girl with neonatal hyperinsulinism. Since congenital hypoglycemia secondary to hyperinsulinism can be a feature of Kabuki Syndrome 1, in-depth phenotyping to detect disease-associated features was recommended by the MDT^19^. However, no Kabuki-like features were evident, thus calling into question a disease association for the identified variant. The second case was a 2-year-old boy with mild motor delay and muscular hypotonia who was included in TRANSLATE-NAMSE due to the suspicion of a neuromuscular disorder. Similarly, no Kabuki Syndrome-associated features were observed.

A functional assessment of the variants was then performed. In both cases, DNA methylation was analyzed using EPIC arrays, as described elsewhere^19^. This was also performed for n= 5 positive (i.e., confirmed diagnosis of Kabuki syndrome) and n= 97 controls (having other diagnoses or VUS in other genes than *KMT2D*). First, CpG sites that showed a significant epigenome-wide association with Kabuki syndrome were identified via an epigenome-wide association analysis. Second, a support vector machine (SVM) was employed for the epi-signature to distinguish between variants with a Kabuki-like methylation pattern and those without. Interestingly, both cases yielded an SVM score of <0,3 indicating that in general, the variants have no effect on DNA methylation. Based on these findings, the variants were reclassified as VUS in accordance with the ACMG criterion “BS3: Well-established *in vitro* or *in vivo* functional studies show no damaging effect on protein function or splicing”. Of note, Supplemental Figure 10 displays an independent case with an SVM score >0.3 but < 0.5. This case was found to have a pathogenic *KMT2D* variant with high-grade mosaicism in blood DNA.

*MDH2* (functional assays: proteomics)

In a 4-year-old girl (case 1127) from a non-consanguineous family who presented with global developmental delay and muscular hypotonia, compound heterozygous VUS c.755C>T, p.Ala252Val and c.884G>T, p.Gly295Val were identified in *MDH2* (NM_005918.2). Additional clinical findings comprised ataxia, stereotypic movements, and growth delay. Serial magnetic resonance imaging studies revealed progressive cerebellar atrophy and delayed myelination. An electroencephalogram revealed frontocentral spikes, although no history of seizures was reported.

The variants in *MDH2* were classified as VUS. Subsequent RNA sequencing from fibroblast RNA revealed no reduction of *MDH2* transcripts and no aberrant splicing. However, subsequent proteomics showed a significant (p=2.45E-09) protein reduction of 60% (log2fold change = -1.316, Supplemental Figure 11A). The protein levels found in the present case were lower than those found in 98 fibroblast proteome samples that were processed in parallel (Supplemental Figure 11B). To date, only three unrelated patients with MDH2 deficiency have been reported in the literature, all of whom presented with a more severe phenotype than the present case^20^. A discussion of molecular and clinical findings within the respective MDT concluded that the variants in *MDH2* were the most likely cause for the encephalopathy in this case, albeit with an attenuated phenotype in comparison to published cases.

*RNASEH2A* (therapeutic consequences in Aicardi-Goutieres syndrome 4)

A 10-month-old girl (case 892) presented with developmental regression, muscular hypotonia, and an inability to maintain visual fixation. Trio ES revealed a homozygous missense variant in *RNASEH2A* (NM_006397.2:c.859T>C, p.Tyr287His), which was classified as a VUS in accordance with current ACMG criteria. Subsequent reverse phenotyping generated laboratory findings highly suggestive of “Aicardi-Goutieres syndrome 4” (positive interferon signature in peripheral blood, increased interferon-alpha activity in the cerebrospinal fluid (CSF), CSF leukocytosis) leading to the reclassification of the variant as “likely pathogenic”. Treatment with the JAK inhibitor Ruxolitinib, which is a proposed treatment option for selected interferonopathies, was initiated. This case illustrates the value of specific laboratory constellations for variant (re-)classification in selected disorders, and the therapeutic consequences of a genetic diagnosis.

### Supplemental Figures

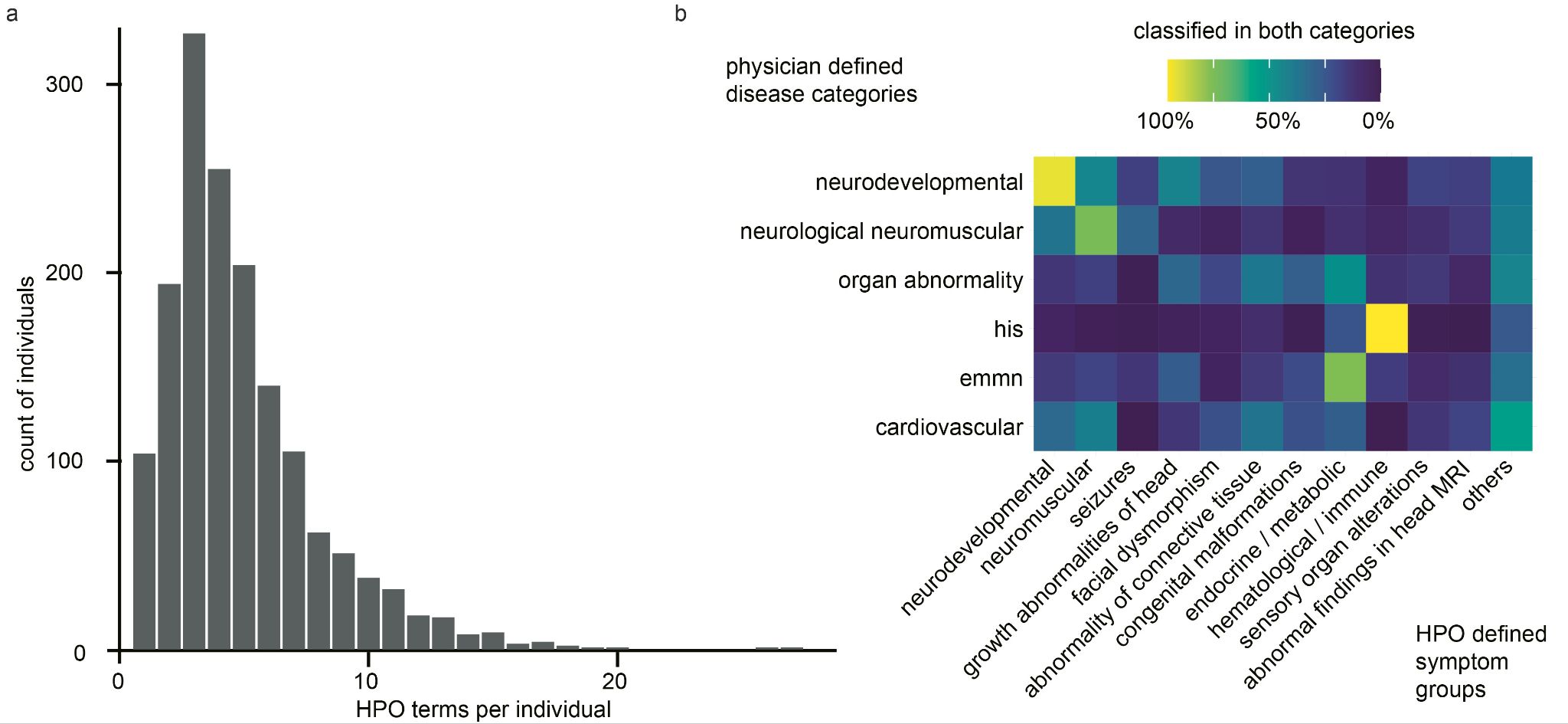

**Supplemental Figure 1: HPO terms and correlation with disease groups.** a) The mean number of Human Phenotype Ontology (HPO)-terms used to describe a patient was 5. b) Each HPO term was also assigned to a higher-order HPO group in order to study the correlation between the higher-order HPO group and the physician-reported disease groups. A high proportion of individuals with neurodevelopmental disorders also had neurological and neuromuscular abnormalities and vice versa.

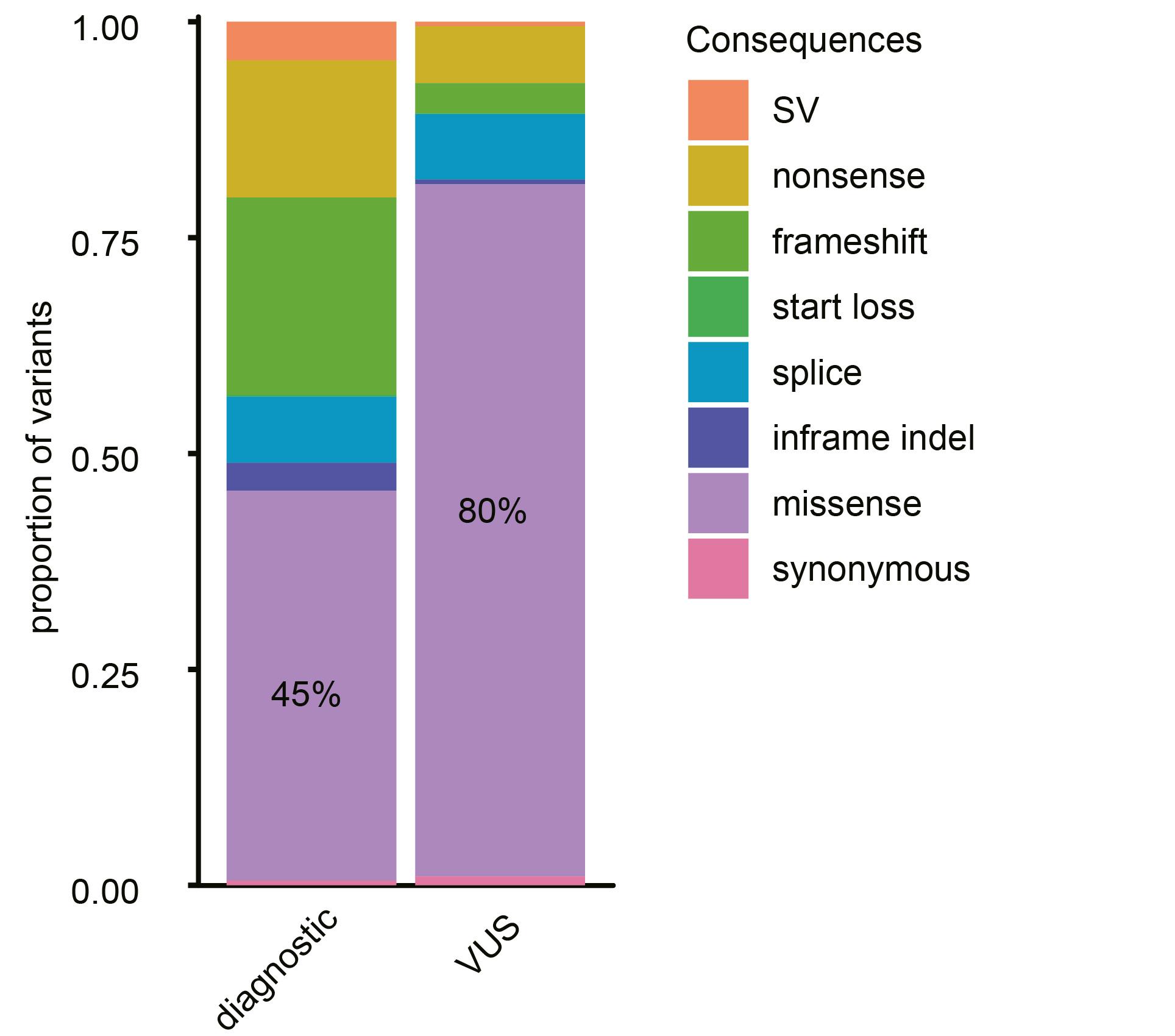

**Supplemental Figure 2: Types of diagnostic variants versus types of variants of unknown significance (VUS).** VUS were enriched for missense variants (80% vs. 45%, p<0.001). SV: Structural variant; indel: insertion or deletion.

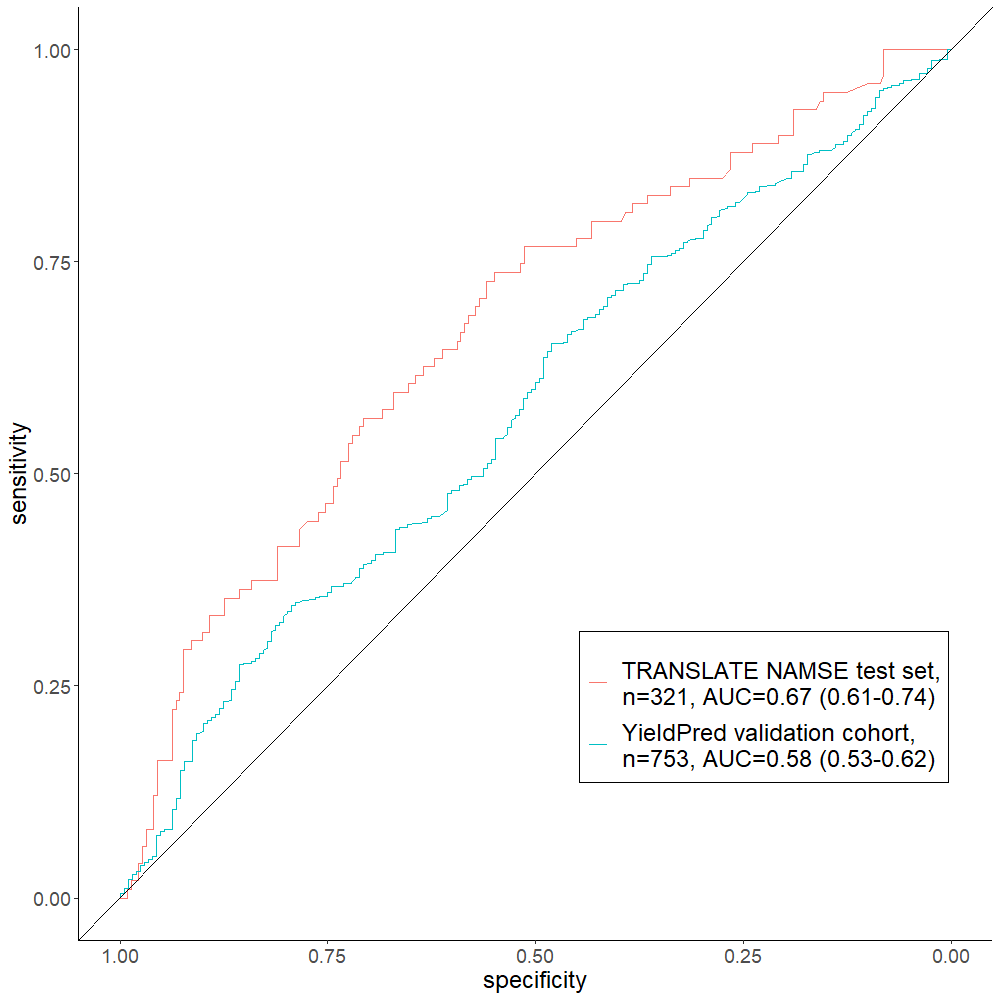

**Supplemental Figure 3: Receiver Operator Characteristics (ROC) curve for Predictor of diagnostic yield on a TRANSLATE-NAMSE test set and on the YieldPred validation set** (Supplemental Table 4). YieldPred could discriminate between solved and unsolved cases in both cohorts. AUC: Area under the curve.

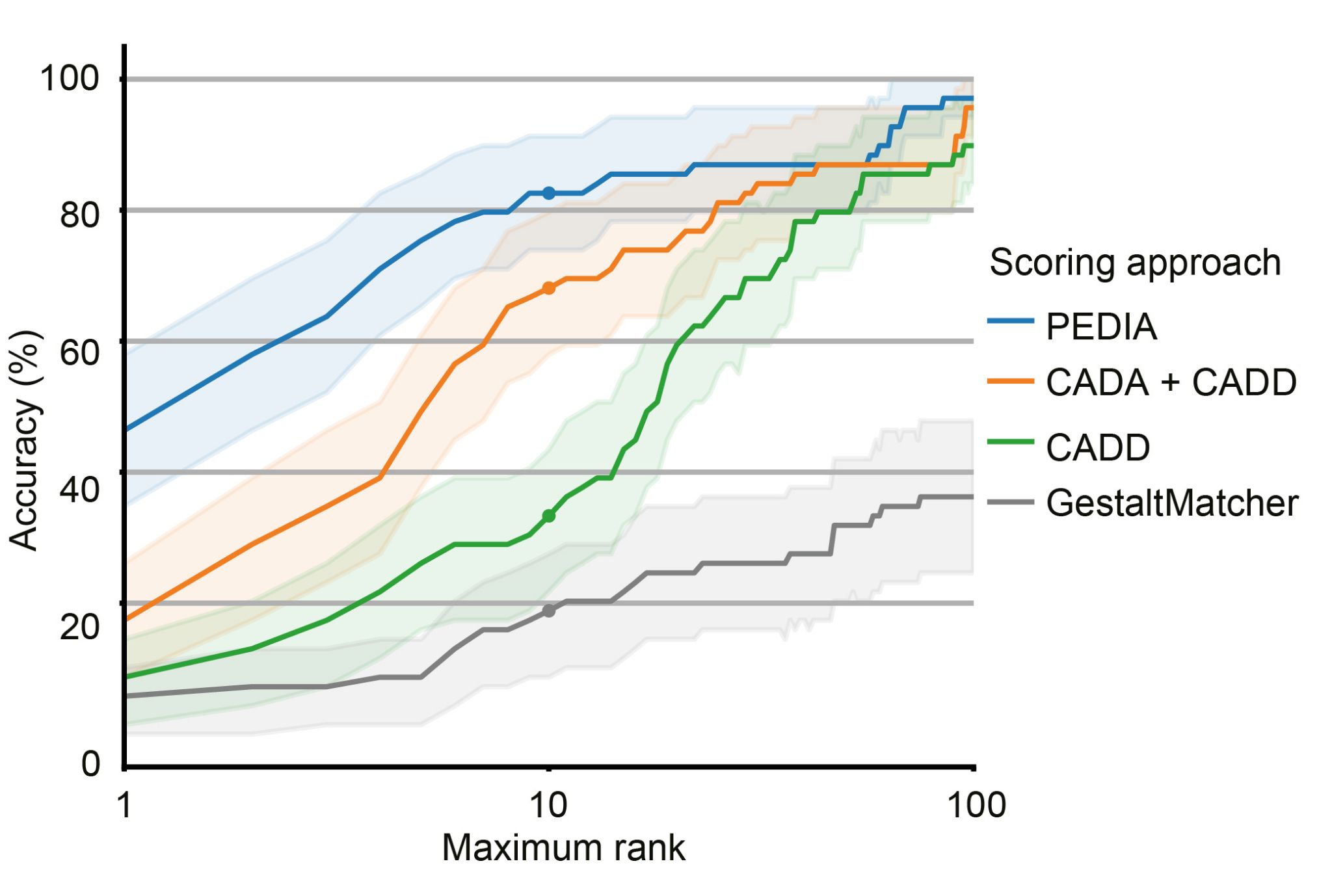

**Supplemental Figure 4:** **Performance of selected variant prioritization approaches.** The validation cohort was used to compare the performance of selected variant prioritization approaches. All disease-associated genes were ranked using the respective variant prioritization method. Subsequently, the proportion of cases detected with the correct disease-associated gene (sensitivity) was shown as a function of the number of disease-associated genes considered, beginning at the top score. The following three approaches for variant prioritization were then tested in solved cases from the PEDIA validation cohort (n=69): 1) only a molecular pathogenicity score (CADD) with a top-10 accuracy of 33%; 2) a feature-based score (CADA) in addition to CADD with a top-10 accuracy of 68%; 3) A gestalt score from facial image analysis (GestaltMatcher) in addition to both CADD and CADA – termed the PEDIA score - with a top-10 accuracy of 83%. Note that bootstrapped 95% confidence intervals are indicated by the lighter shading around the lines. CADA: Case Annotation and Disorder Annotation; CADD: Combined Annotation-Dependent Depletion; PEDIA: Prioritization of Exome Data by Image Analysis

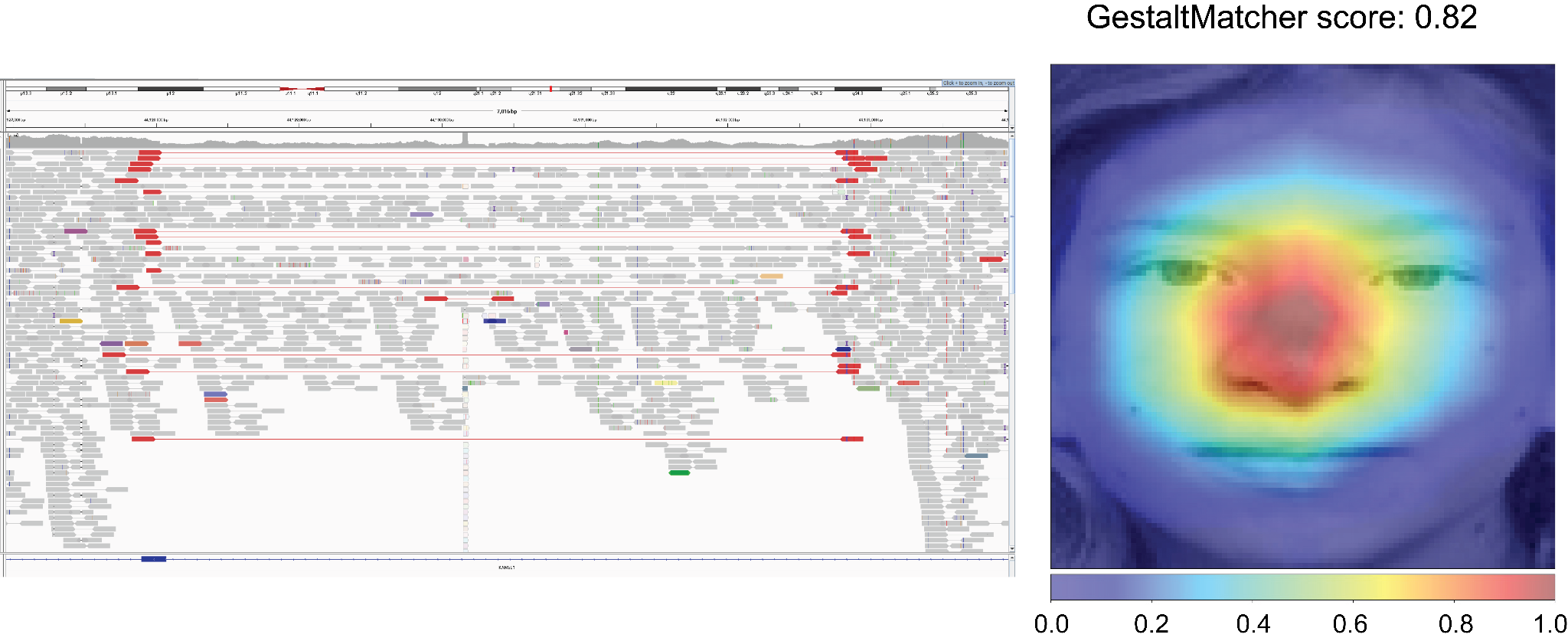

**Supplemental Figure 5: Value of next-generation phenotyping.** Facial image analysis of case 393 with GestaltMatcher suggested a high similarity to Koolen-De-Vries syndrome. After inconclusive exome sequencing, analyses targeted at *KANSL1* and genome sequencing were performed. These revealed a causal 4.7 kb *de novo* deletion in *KANSL1**^16^*

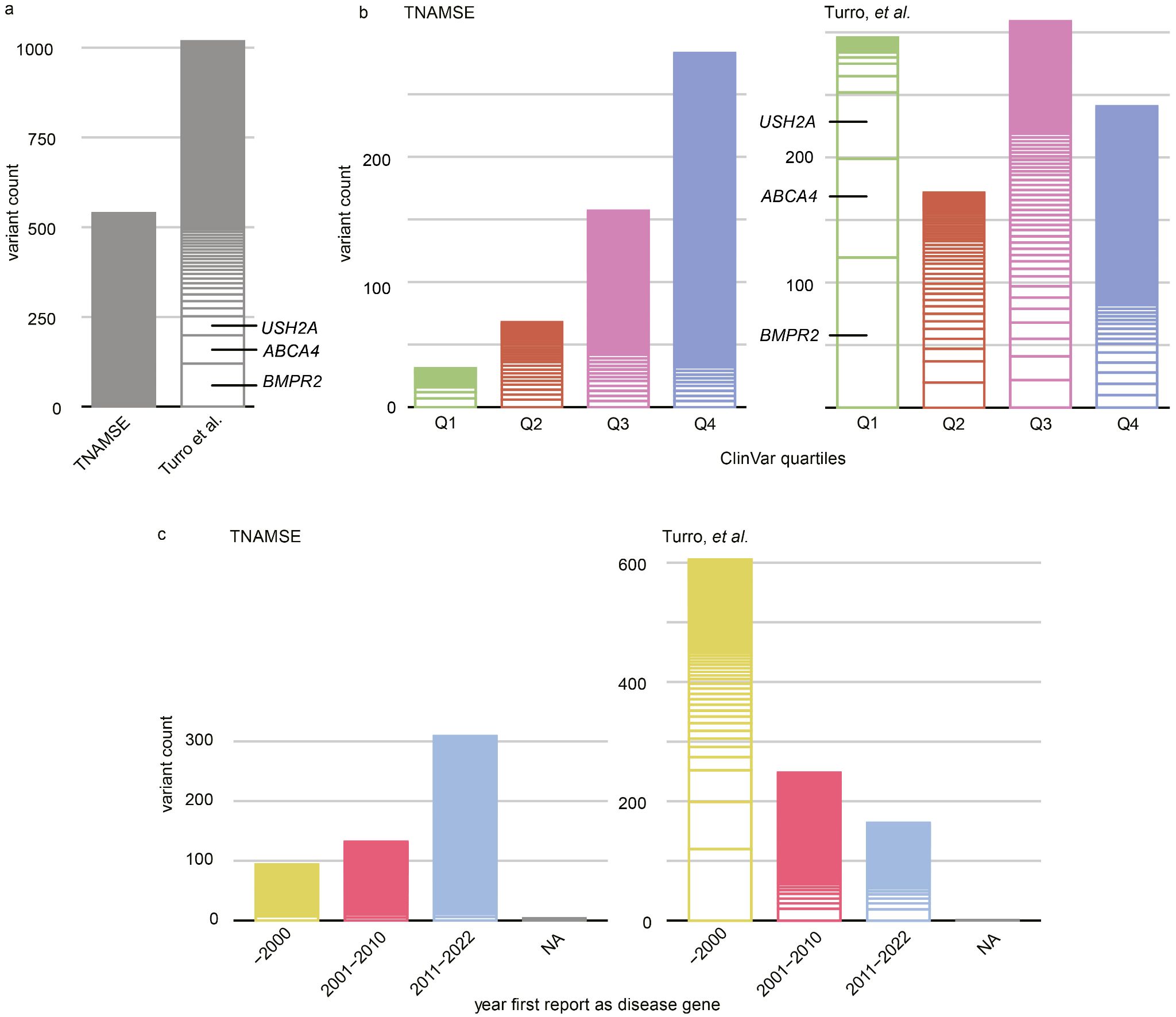

**Supplemental Figure 6: Comparison of molecular diagnoses in the rare disease programs of two different national health systems (TRANSLATE NAMSE Germany, and Turro, et al. UK**^21^**).** A) The total number of (likely) pathogenic variants reported by Turro, et al. was larger. However, many of these were in DGGs that are subject in Germany to tests such as panel testing. B) Disease-associated genes were ranked according to the number of (likely) pathogenic variants submitted to ClinVar, with the most frequently affected genes in the 1st quartile and the least frequently affected genes in the 4th. The majority of (likely) pathogenic variants reported in TRANSLATE NAMSE were in disease-associated genes from the 4th quartile. C) The causative disease-associated genes found in TRANSLATE NAMSE and by Turro, et al. were sorted according to the year in which they were first reported as disease-associated genes. The different distributions illustrate the high proportion of recently described disease-associated genes in the TRANSLATE NAMSE ES cohort. NA corresponds to genes that did not reach disease-associated gene status prior to the publication of the present study.

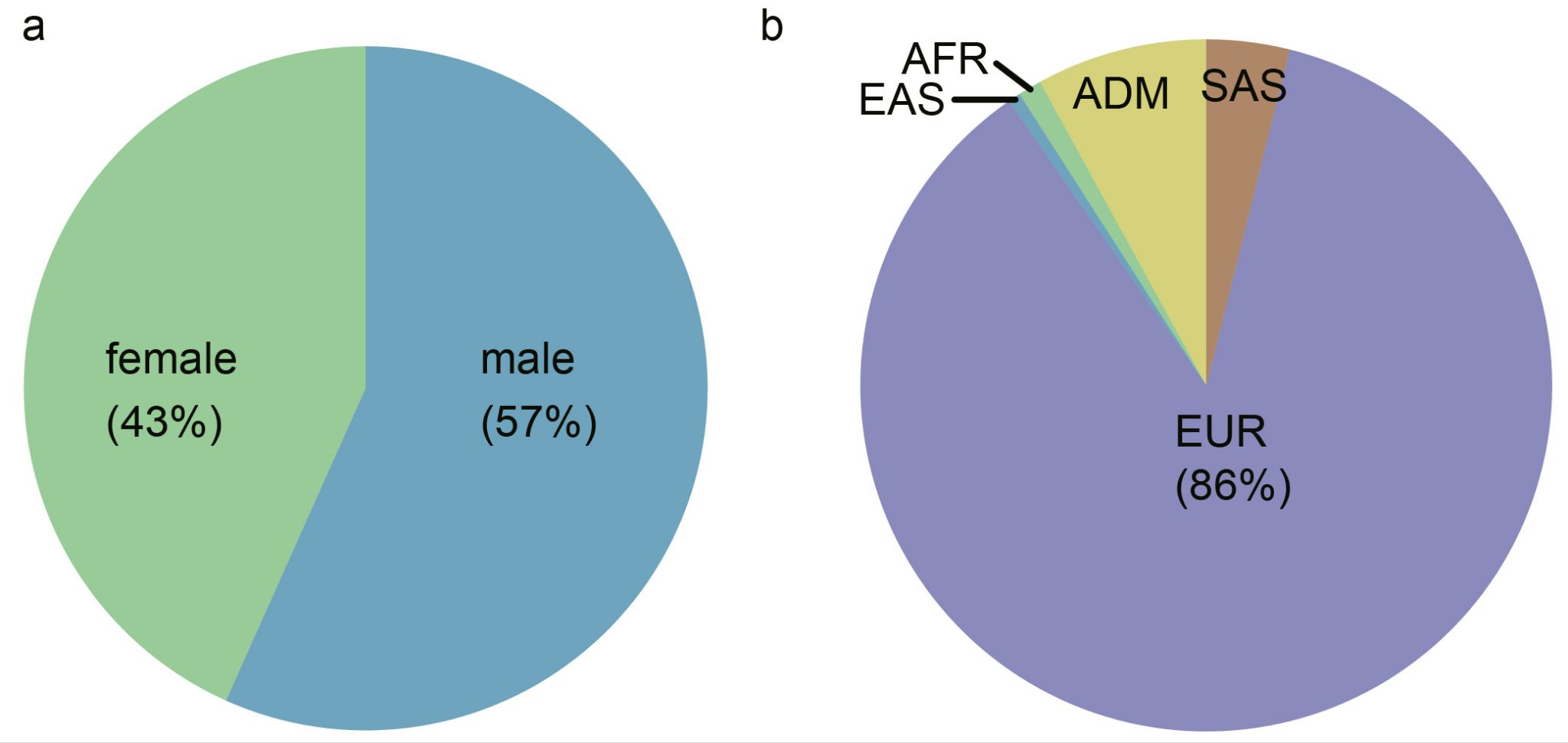

**Supplemental Figure 7**: **Sex and population background of the TRANSLATE NAMSE exome sequencing (ES) cohort.** Pie chart showing (a) the sex of 1,577 individuals from the TRANSLATE NAMSE ES cohort and (b) the available population background of 1,365 individuals. As expected, the majority of individuals (86% or 1,180) were assigned to the European population. EAS: East Asian; AFR: African; ADM: admixed; SAS: South Asian; EUR: European.

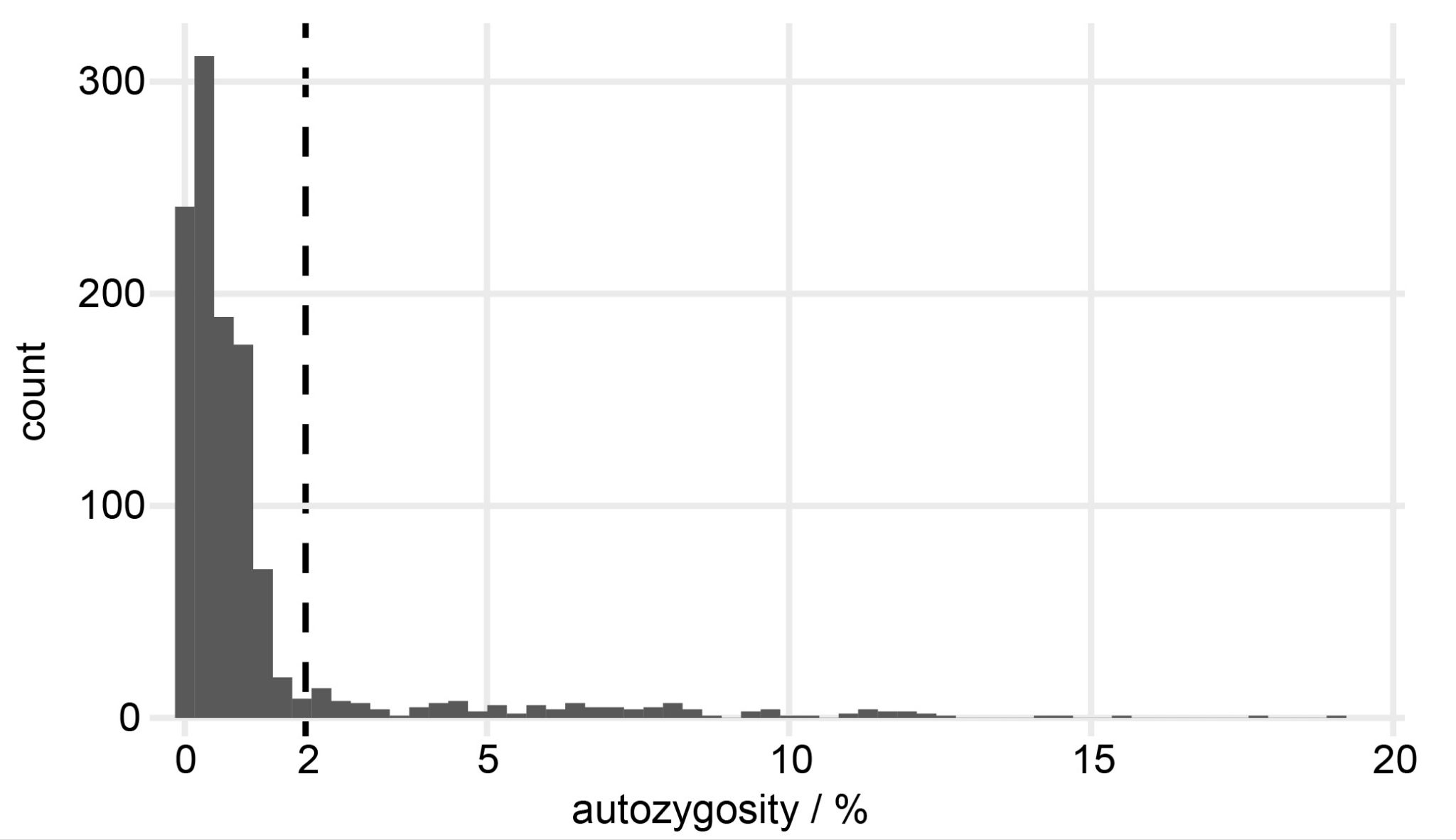

**Supplemental Figure 8**: **Histogram showing the autozygosity distribution** in the subcohort of n=1,158 individuals for whom autozygosity data were obtained. The x-axis represents the autozygosity, and the y-axis represents the count of individuals with the respective autozygosity. The vertical line indicates the 2% cut-off used to classify individuals as having low or high autozygosity.

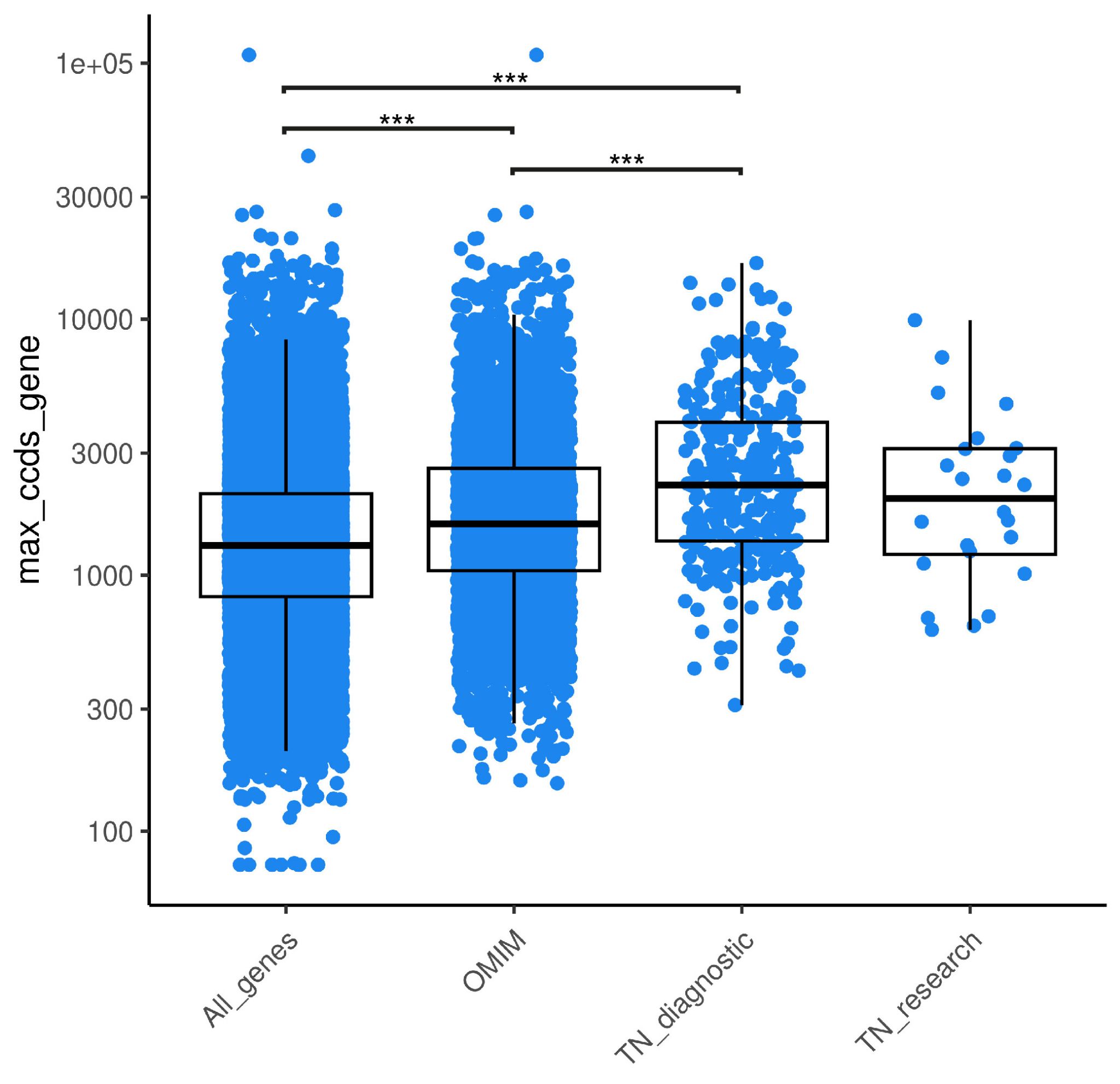

**Supplemental Figure 9:** **Disease genes in TRANSLATE NAMSE cohort are significantly longer than all genes/OMIM genes.** Boxplots of the coding length of different gene sets are shown. The mean coding length of disease-associated genes in the TRANSLATE NAMSE (TN) ES cohort, which was subdivided into known disease-associated genes (TN_diagnostic) and novel disease-associated genes (TN-research), was significantly longer than for all genes or for all OMIM genes (Pairwise t-test. P-values were adjusted by Bonferroni correction. *** = p<0.001).

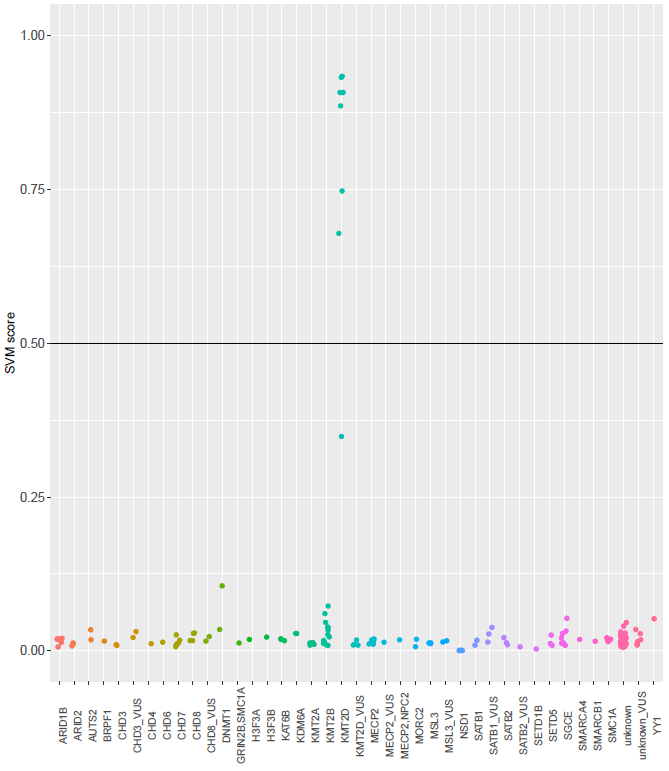

**Supplemental Figure 10:** Monogenic disorders of chromatin regulatory proteins cause characteristic methylation epi-signatures. These patterns could be identified using EPIC-array data and a support vector machine (SVM) that was trained on positive controls for each gene. A score above 0.5 is indicative of a pathogenic variant in the respective gene. Scores of between 0.3 and 0.5 can be caused by mosaicism of pathogenic variants. Individuals with *de novo* variants in *KMT2D* (see Case reports of particular interest), formerly classified as “likely pathogenic” had a SVM score < 0.3 (here displayed as KMT2D_VUS) excluding an effect of these variants on methylation. Thus, variants could be reclassified as VUS.

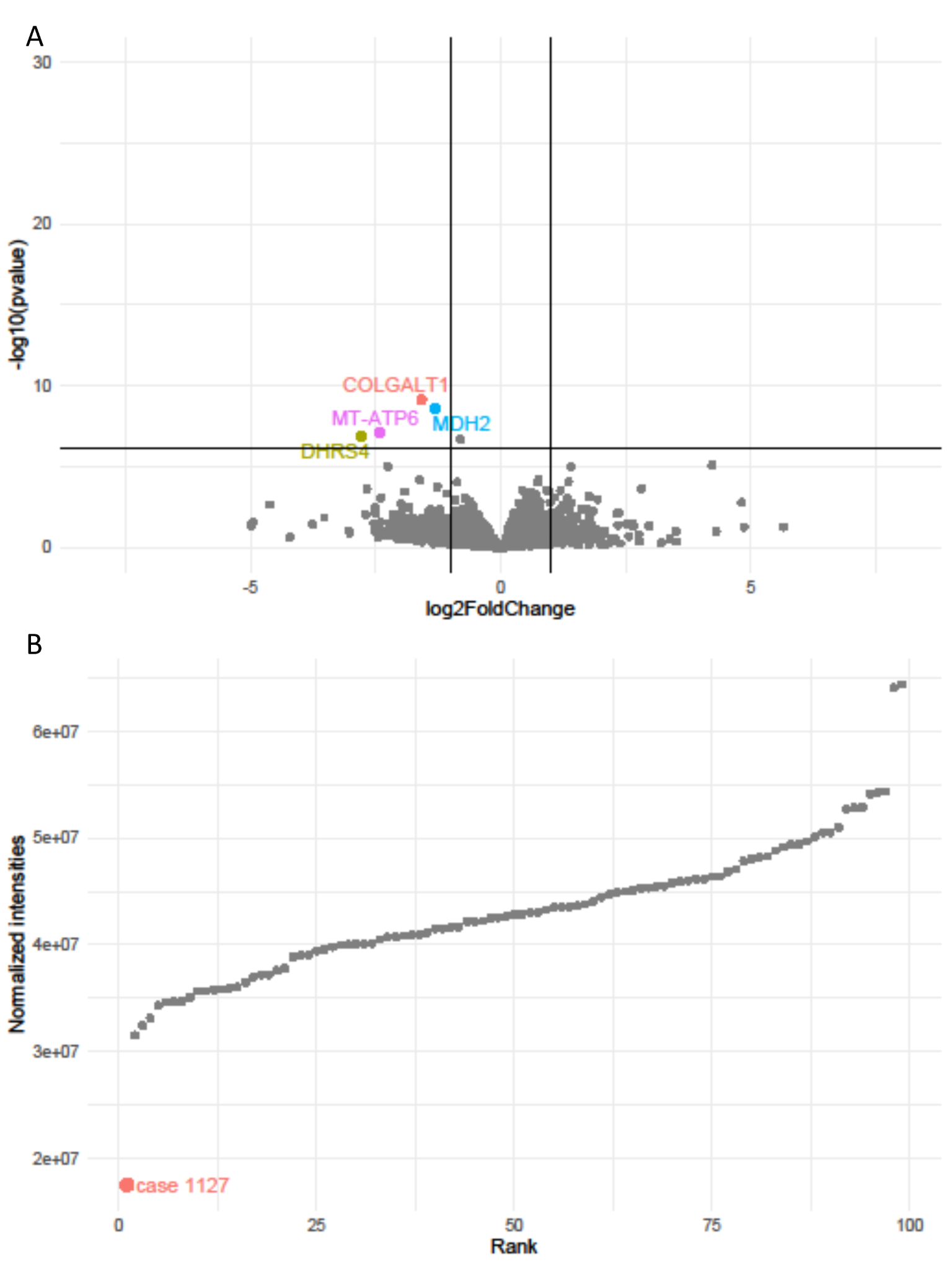

**Supplemental Figure 11: Significant MDH2 reduction in fibroblasts.** Exome sequencing in case 1127 revealed compound-heterozygous missense variants of uncertain clinical significance in *MDH2.* Subsequent proteomics in fibroblasts showed significantly reduced intensities for MDH2 (A) with a log2fold change of -1.316 (p= 2.45E-09). This resulted in a reclassification of the variants to likely pathogenic. Off note, proteomics analysis detected an outlier, LAGE3 (log2fold change -7.116, p= 9.73E-46) which however is a false positive as a rare SNP affects a peptide which is subsequently undetectable by mass spectroscopy. (B) MDH2 had the lowest normalized intensity in proteomics in comparison with 98 controls.

### Supplemental Tables

Supplementary Tables can be found at:

<https://drive.google.com/drive/folders/1AXXDweePuc12SoLYLIsq92XYb0R6eC57>

**Supplemental Table 1:** Complete TRANSLATE NAMSE ES dataset, comprising information on all 1,577 cases included in the present study. Due to its size, the table has been submitted separately.

**Supplemental Table 2:** Dual diagnoses

**Supplemental Table 3:** Candidate genes and novel disease-associated genes

**Supplemental Table 4:** YieldPred validation cohort

**Supplemental Table 5:** PEDIA validation cohort

**Supplemental Table 6:** Secondary findings
