## Supplementary material for "Next-generation phenotyping integrated in a national framework for patients with ultra-rare disorders improves genetic diagnostics and yields new molecular findings": Rebuttal

Bonn, March 2023

Dear Dr. Ranzoni,

We thank you for your careful review of our manuscript (NMED-A118820) and the interest of the editorial team in the results of the ongoing national program for rare diseases in Germany.

We agree with the editors that the context of the study should be explained more clearly, and that more information should be provided on the characteristics of the cohort, the definition of the diseases, and the inclusion/exclusion criteria.

With respect to context, in the original manuscript, we refrained from providing detailed information on current efforts to improve the management of patients with suspected rare disorders within the German healthcare system for reasons of brevity. However, in response to the feedback of the editors, we now provide more detailed information on this issue both immediately below and in the revised manuscript.

In the German healthcare system, new diagnostic approaches, such as exome and genome sequencing, must be evaluated for cost efficiency before they can be integrated into routine care and reimbursed by public health insurance. Evaluation of the cost-efficiency of new molecular genetic diagnostic procedures for rare diseases was thus a major goal of the TRANSLATE NAMSE project. Given that all genetic services in Germany can also be provided by physicians in private practice, an important aim of TRANSLATE NAMSE was to demonstrate that centers of rare diseases (CRDs) can serve as an effective gatekeeper for the most advanced - and usually more expensive - diagnostic tests in order to avoid an explosion in costs. Data from the project have now shown that the CRD concept is economically feasible and can facilitate the diagnosis and management of suspected rare disease patients. These results were published in a German article in Deutsches Ärzteblatt, which is the official journal of the German Medical Association (Bundesärztekammer) and the National Association of Statutory Health Insurance Physicians (Kassenärztliche Bundesvereinigung) (Rillig et al., 2022).

Following this positive evaluation of TRANSLATE NAMSE the concept is now being expanded to include all CRDs and sequencing centers that fulfill specified quality criteria. In the initial 3-year phase of TRANSLATE NAMSE, patient recruitment was performed at 10 CRDs, and a total of four centers were responsible for sequencing. In the second phase of the project, which commenced in 2022, recruitment is being performed at a total of 21 centers, 18 of which are also responsible for sequencing. In addition, further potential recruitment and sequencing sites are currently under review.

In contrast to the work of Rillig, *et al.* that focuses on exome sequencing within the German healthcare system, our manuscript describes the actual molecular findings and the concepts that we consider to be more generalisable.

We are therefore very pleased that AI/machine learning aspects of the present study that are efforts to increase the effectiveness (via YieldPred) and efficiency (via PEDIA) of the diagnostics of ultra-rare diseases have met with interest. Furthermore, we acknowledge that in the original manuscript, the distinction between the use cases of YieldPred and PEDIA was unclear. In the revised manuscript, we have addressed this by presenting clearer explanations of both the rationale for, and the performance of, the two approaches. In addition, we now present the respective results in separate subsections. Briefly, the machine learning tool YieldPred was designed to predict the likelihood that exome sequencing (ES) will lead to a molecular diagnosis in a given patient, as based on the respective clinical features only. YieldPred was designed using the results of a multivariate regression analysis with Lasso statistics, which was performed on phenotypic data from the entire TRANSLATE NAMSE ES cohort (n= 1,577 cases), following annotation with HPO terms. In the revised manuscript, we explain that this model has now been made available as a web-service.

With regards to diagnostic efficiency, we demonstrated that the analysis of facial portraits using the AI tool PEDIA (Prioritization of Exome Data by Image Analysis, PEDIA) can facilitate variant prioritization for ultra-rare diseases. This compandion study required additional informed consent for portrait analysis and included a subset of 224 individuals.

The revised manuscript refers to these two approaches collectively as next-generation phenotyping (NGP), an acronym that is increasingly used for these types of analysis in the bioinformatic literature.

Since the submission of the original manuscript, and the expansion of the TRANSLATE NAMSE project to further CRDs, we have curated NGP validation cohorts. These comprise an additional 753 patients for YieldPred and 149 patients for PEDIA. In addition, we now provide open-source code repositories and FAIR databases for all of our software - including the training data - in order to allow the international bioinformatics community to integrate YieldPred and PEDIA into workflows for rare disease diagnostics, and to customize them as required. This information is now presented in the revised manuscript.

Once again, we thank the editor and reviewers for their thorough evaluation of our work and for their constructive comments, which have allowed us to improve the quality of our manuscript. We look forward to your response, and hope that our work will now be considered suitable for publication. Below, we provide point-by-point responses to the comments and queries of the reviewers.

**Reviewer #1**: The authors say on page 1: “With the entire cohort data, we developed a tool to predict the diagnostic yield from the clinical features of a patient if advanced molecular testing strategies are applied.”

What do you mean by “the entire cohort data”? The subcohort of 211 individuals? This could be clearer. Could you also please be more precise on which data the algorithm was trained on?

**Response**: We thank the reviewer for this comment. In the revised manuscript, we now differentiate more clearly between the two machine learning models that were applied in the present study. The aim of the first approach was to improve diagnostic effectiveness for ultra-rare disorders by developing a novel machine learning tool on the basis of a multivariate

regression analysis with Lasso statistics. This model was designed to predict the likelihood that ES would lead to a molecular diagnosis in a given patient, as based on the respective clinical features only. This model is referred to in the revised manuscript as YieldPred. The aim of the second approach was to improve diagnostic efficiency for ultra-rare disorders, and involved application of the previously established AI tool PEDIA (Prioritizing variants in Exome Data with results from Image Analysis). For YieldPred, the data of the entire TRANSLATE NAMSE ES cohort (n= 1,577 patients) was used, since all individuals had been deeply phenotyped with HPO terms. PEDIA involved the analysis of facial portraits with the AI tool GestaltMatcher, and required separate informed consent, which was only available for a subset of patients. In the original manuscript, this consent was provided by 211 of the 1,577 patients from the TRANSLATE NAMES ES cohort. Since the submission of the original manuscript, the consent of an additional 13 TRANSLATE NAME study participants has been received for the AI/machine learning analyses, and their data are now reported in the revised manuscript document.

Since the submission of the original manuscript, we have also curated validation cohorts comprising 753 individuals for YieldPred and 149 for PEDIA from newly included TRANSLATE NAMSE CRDs. Analyses of data from these validation cohorts confirm our initial results. All of these validation data are available as Supplemental Material and via GestaltMatcher DB. The latter is a FAIR database, which requires access control to comply with GDPR: A registration is possible with the following invitation link:

<https://db.gestaltmatcher.org/users/sign_up?token=ffBou7UQrHufMoHXm2hu>

**Reviewer #1**: Do you have validation data? Why do you think that the AI model works? Could you say a little bit more about the sensitivity/specificity of the system?

**Response**: We thank the reviewer for this important comment. To clarify, our data was split into separate training, validation, and test sets, and this is now specified in the revised manuscript. The respective text on page 5 reads :

**Predicting the diagnostic yield of ES using machine learning (YieldPred).** Analyses were then conducted to investigate whether the phenotype predicted the diagnostic yield of ES. For this purpose, a least absolute shrinkage and selection operator (LASSO) analysis for binary outcomes was performed. To reduce the phenotypic dimension and to increase interpretability, HPO terms were first aggregated into 49 non-overlapping phenotypic groups. These phenotypic groups were used as predictors in the LASSO analysis. The resulting model was able to discriminate between solved and unsolved cases (Supplemental Figure 3, area under the curve (AUC)=0.67, 95%-confidence interval=0.61-0.74, on a held-out test set of the ES cohort, n=321), and yielded the HPO groups “dysfunction of higher cognitive abilities”, “hematological abnormalities”, and “ataxia” as very influential predictors in terms of the establishment of a molecular diagnosis via ES (Figure 5a). This model was then used to build the machine learning tool YieldPred, which was validated using an external cohort (Supplemental Figure 3, n=753, AUC=0.58). YieldPred has now been made available as a web service that can be used to estimate the diagnostic yield of ES on the basis of the phenotypic features of a given patient ([https://translate-namse.de/](http://tnamse.de/)).

**Reviewer #1**: What is with the race/ethnicity and gender [sex] breakdown?

**Response**: Sex and population background were inferred using peddy (Pedersen & Quinlan, 2017). This is now stated in the case information in the Online Methods. The female-to-male ratio was 8:10. The majority of individuals were of European ethnicity, which is representative of the population affected by rare disorders in Germany.

**Reviewer #1**: I would also like to better understand what the AI model [PEDIA] exactly does (more information than just “to analyze medical imaging and to interpret genetic variants”.)

**Response**: The PEDIA approach was first described by Hsieh, *et al.* in 2019. PEDIA was designed as a prioritiztion algorithm, and is an AI driven approach that uses portrait photographs for the interpretation of clinical exome data. The design of PEDIA is based on the fact that since multiple monogenic disorders are characterized by facial dysmorphism, facial analysis by an AI tool trained in all known facial dysmorphism-associated syndomes may expedite the clinical dignosis of a genetic syndrome and facilitate the otherwise time and energy intensive analysis of genetic data. The main difference between PEDIA and standard prioritization algorithms, such as e.g. Exomiser is the additional use of a syndromic similarity score, which is provided by the AI tool GestaltMatcher, as first described by Hsieh, *et al.* in 2022. The gestalt score can be used to quantify the phenotypic match of a patient with a disorder which is also considered in variant classification according to the ACMG guidelines. The deep convolutional neural network (DCNN) of GestaltMatcher is updated as new data become available, and/or new algorithms outperform the old model. The present study applied the latest performance of GestaltMatcher, which is described in detail elsewhere (Hustinx et al., 2022).

From a computer vision perspective, building a good classifier and deducing the facial features that are most informative for the machine (explainable AI) are two different disciplines. For the further development of NGP into certifiable medical products, certain AI software requirements must be met. One such requirement is explaining the AI results in such a way that physicians can reason about the decision of the model. To address this requirement, we applied Gradient-weighted Class Activation Mapping (Grad-CAM) to produce a coarse localization map, which highlights regions in the portrait photographs that were important in terms of disease classification (Selvaraju et al., 2016) Grad-CAM uses the gradients flowing into the final convolutional layer to generate a heat map for any syndrome that GestaltMatcher has learned. In the example shown below, which is also described in the supplement, we overlaid the heat map that was generated for Koolen de Vries syndrome on the original image of the patient. As for most syndromologists, the AI GestaltMatcher rated a high nasal bridge with a bulbous nasal tip as the most informative feature in terms of establishing the diagnosis.

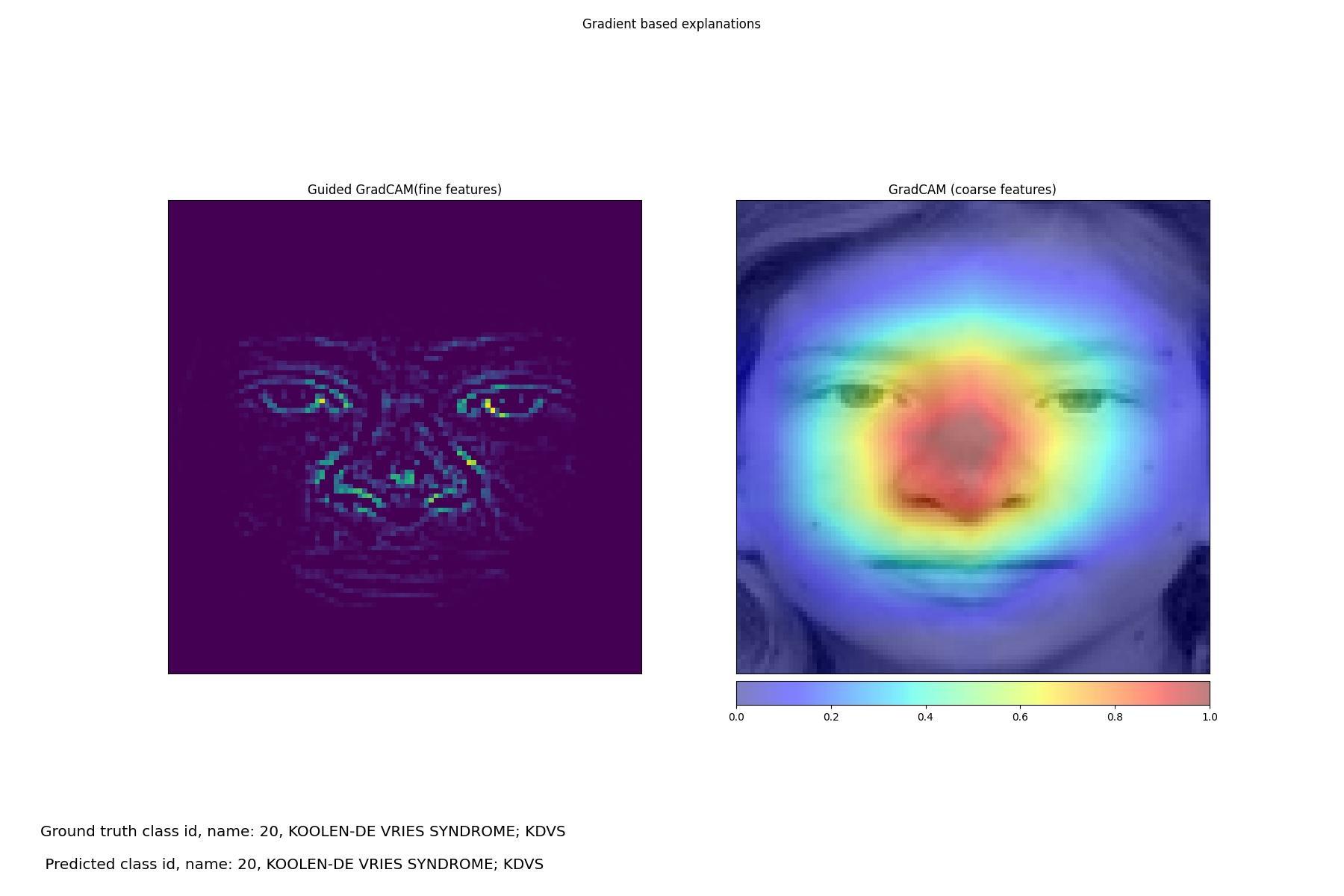

**Reviewer #1**: Why did 211 individuals consent to the analysis of the portrait photos by the AI tool? What are the reasons why individuals did not consent? If the authors do not want to include all of the above information in the main manuscript, this information should be at least included in a supplement.

**Response**: We thank the reviewer for this comment, and agree that these are important issues. Facial portraits and consent to their analysis via AI were obtained in a separate, companion study “PEDIA” to TRANSLATE NAMSE. Although all 10 of the original TRANSLATE NAMSE recruitment sites also had IRB approval for computer-assisted image analysis, only two sites did in fact recruit patients for the PEDIA analyses. In total, 473 patients were informed about the PEDIA study, and 211 agreed to participate (47%).

No data are available to us concerning the specific reasons why these 473 individuals either refused or agreed to participate in these analyses. Interestingly, however, a large proportion of the patients who agreed to participate had some form of facial dysmorphism. We hypothesize that patients who are of the opinion that they have some form of facial dysmorphia may rate the potential value of PEDIA more highly. We plan to investigate this issue in a follow-up study. At writing, no reliable estimates on the prevalence of facial dysmorphism in patients with genetic diseases are available which is partly due to the fact that there are no generally accepted measurement methods for it. However, for roughly one-third of all OMIM entries, the clinical synopsis lists an abnormality of the face (HP:0000271) or a more specific term. The issue of whether computer-assisted pattern recognition is more sensitive than a clinical assessment by a dysmorphologist remains unclear. The fact that individuals without facial dysmorphism also participated in the study is therefore valuable.

**Reviewer #1**: The authors say: “A clear separation between diagnostics and research cannot and should not be applied for patients with ultra-rare disorders.”

It would be helpful to clarify why. I assume that the authors forgot to incorporate this (see the sentence in German at the bottom of the page that would also need to be deleted).

**Response**: We thank the reviewer for these comments. This issue of the separation between diagnostics and research was also raised by Reviewer #3. In the original manuscript, the phrasing was inaccurate and we apologize for the resulting confusion. To clarify, we intended

to refer to the tertiary clinical setting, i.e., as opposed to the primary care setting, rather than to the research setting. We agree with Reviewer #3 that the separation between diagnostics and research cannot, and should not, be eliminated, and have now reformulated this section of text accordingly.

The revised text reads: “[...] Therefore in the tertiary, academic setting, collaboration between experts from diagnostics and research is highly relevant for patients with suspected ultra-rare diseases and a lack of definitive diagnostic findings [...]”

We apologize for the inadvertent retention of a German language sentence, and thank the reviewer for pointing this out. This sentence has now been deleted.

**Reviewer #2**: The authors proposed a web-based tool to predict the diagnostic yield from the clinical features of a patient if advanced molecular testing strategies are applied. Given the scarcity of genetic testing due to high cost and infrastructure requirements, such a tool may speed up the rare diagnosis and properly channel healthcare resources.

However, the overall writing style of the paper is quite confusing and needs a thorough proof-check. The novelty of the work should be clearly highlighted in the 'Introduction'.

The core of the manuscript is the development of a simple LASSO regression model with 49 clinical features. But the choice of model selection is not clear to me given that the authors formulated it as a classification, not a regression problem.

**Response**: We thank the reviewer for these important comments, and agree that sections of the manuscript text required reformulation. This has now been done, and the revised manuscript has been proof-read by a native English speaker. We thank the reviewer for pointing out that the choice of model selection was unclear. To clarify, the problem was indeed a classification issue (case solved or unsolved), which was addressed via a logistic regression approach for binary responses. This is now described in a separate paragraph in the results section:

**Predicting the diagnostic yield of ES using machine learning (YieldPred).** Analyses were then conducted to investigate whether the phenotype predicted the diagnostic yield of ES. For this purpose, a least absolute shrinkage and selection operator (LASSO) analysis for binary outcomes was performed. To reduce the phenotypic dimension and to increase interpretability, HPO terms were first aggregated into 49 non-overlapping phenotypic groups. These phenotypic groups were used as predictors in the LASSO analysis. The resulting model was able to discriminate between solved and unsolved cases (Supplemental Figure 3, area under the curve (AUC)=0.67, 95%-confidence interval=0.61-0.74, on a held-out test set of the ES cohort, n=321), and yielded the HPO groups “dysfunction of higher cognitive abilities”, “hematological abnormalities”, and “ataxia” as very influential predictors in terms of the establishment of a molecular diagnosis via ES (Figure 5a). This model was then used to build the machine learning tool YieldPred, which was validated using an external cohort (Supplemental Figure 3, n=753, AUC=0.58). YieldPred has now been made available as a web service that can be used to estimate the diagnostic yield of ES on the basis of the phenotypic features of a given patient ([https://translate-namse.de/](http://tnamse.de/)).

**Reviewer #2**: The study cohort should be clearly described in a patient characteristics table and model training and validation strategy should be highlighted. The discussion in the Result section with specific subgroups is not comprehensible as the training and test split is not described. The study finding(s!) remains unclear from the discussion. The performance of the LASSO fit is never mentioned in the manuscript.

**Response:** We thank the reviewer for these comments, and agree that the multiple findings of the study must be presented using a clearer structure. Each patient from the cohort is now described in Supplementary Table 1 using HPO terms. Patients who consented to the sharing of their portrait image can be found in the GestaltMatcher database with the case ID, which is also listed in Supplementary Table 1. To protect sensitive data and comply with GDPR, the use of this resource requires registration, which is possible with the following link:

<https://db.gestaltmatcher.org/users/sign_up?token=ffBou7UQrHufMoHXm2hu>

The differing objectives of the two machine learning approaches are now clearly stated in the Introduction, and their results are presented in separate paragraphs. Lasso was used to analyze the effectiveness of ES, and the resulting model can also be used to estimate the likelihood that ES will result in a molecular diagnosis in a given patient, as based on the respective HPO terms only. For this reason, we now refer to this tool as YieldPred (i.e., Yield predictor). We also evaluated the ability of the previously established AI prioritization algorithm PEDIA to improve diagnostic efficiency for ultra-rare disorders. This tool makes use of a syndromic similarity score, which is generated via the analysis of a facial portrait.

**Reviewer #3:** [The authors] have defined a large, heterogeneous, evolving German program to evaluate undiagnosed children [and adults]. In a way, this seems to be the German answer to the U.S. undiagnosed diseases network – but oddly, that program is never mentioned here.

- Given that this program is situated in a country with a specific healthcare reimbursement scheme, what is the generalizability of this program? The hallmark of scientific papers is generalizability and utility to others.
- It is not clear to me that what was done here is relevant to other countries and health care systems. The overall project seems to be a work in progress – as it no doubt needs to be to best serve the patients and take advantage of the continuously evolving technology. However, this makes it a weaker scientific paper – it is just not a study or a research project. In the abstract, they define it as a ‘study’ but this is a bit of a stretch as it seems more like a program with the paper being a description of the program.

**Response:** We thank the reviewer for these important comments. The stated goal of the International Rare Disease Research Consortium is that by 2027, all patients who come to medical attention with a suspected rare disease should be diagnosed within one year, if the respective disorder has been described in the medical literature. We do, of course, acknowledge that many countries are trying to meet this goal and are therefore in the process of attempting to improve the effectiveness and efficiency of healthcare for rare diseases.

In our work we were analyzing scientific questions of relevance to the international rare disease community, and not issues of specific relevance to the German public health system only. Issues relating to the cost-effectiveness of TRANSLATE NAMSE were addresed by Rillig, *et al.* and were not a focus of the present analyses.

Besides novel molecular findings, the present study also evaluated the potential of two machine-learning approaches (i.e., YieldPred and PEDIA) to facilitate diagnostics in the field of rare diseases. The results demonstrate their value in increasing diagnostic effectiveness and efficiency in a field for which the management of large and diverse data volumes is a major challenge involving time-consuming and expensive procedures. We will continue to update these tools in order to increase their performance. A key feature of the algorithms is that they can be applied in any healthcare system around the world, since they are based on internationally agreed clinical criteria. The issues of reimbursement and cost-effectiveness are beyond the scope of the present analyses, whose focus was on the analysis of phenotypic and genetic data and their relevance for the diagnosis and management of rare disease.

**Reviewer #3:** There are innumerable undefined terms in the paper that make the claims difficult to evaluate. For example, in this text, there are four undefined terms: “This system has proven effective for some of the #more common rare diseases# and the ones that are caused by variants in #well-defined molecular pathways# (Supplemental Material). However, in total, these disorders only represent a #small subset of all rare disorders#. For #highly heterogeneous phenotypes#. Item 4 titled “Supplementary materials” does not include a listing of “well-defined molecular pathways“, though I doubt such a thing can be defined – there is no consensus definition of such pathways. My point here is not to demand strict definitions of all these terms but rather to highlight the pointless nature of this purple prose. Since none of these terms are defined what does any of this actually mean? What this seems to be is an effort to make a program sound like a research study where hypotheses are tested, but it just isn’t the case.

**Response:** We thank the reviewer for these important comments, and revised the manuscript with a native English speaker. The large number of diagnoses established in the present cohort suggests that each specific phenotype had a very low prevalence. To clarify this, the results were compared with the relative number of (likely) pathogenic ClinVar submissions for the respective genes. Genes can then be ordered accordingly and result in an assignment to quantiles. Consequently, each quantile also defines a set of genes. Since the order we set corresponds to decreasing incidence, we now refer to disorders caused by (likely) pathogenic variants in genes of the first quartile as “more frequently identified rare diseases” instead of #more common rare diseases#.

We agree with the reviewer that the term #well-defined molecular pathway# is non-specific, and in the revised manuscript, this term is not used in either the main manuscript or the Supplemental Materials.

The phrase #highly heterogeneous phenotypes#, is for instance also used in the work of Kochinke, *et al.* which is a systematic analysis of the high [genetic] heterogeneity of intellectual disability (Kochinke et al. 2016). This is no excuse but perhaps an explanation why we used it. We now rephrased to “phenotypes with high genetic heterogeneity”.

The TRANSLATE-NAMSE study ran over a period of three years. At the same time, the PEDIA protocol was also studied prospectively for the first time. In this work we report about the molecular findings and validate the AI related results on data that was acquired after the study ended.

**Reviewer #3:** It makes no sense to evaluate the “diagnostic yield of ES”. This is entirely dependent on the robustness of the evaluations upstream of this study. And, paradoxically, the better those are - the worse the yield. I note in the vignettes that some had NGS panel and microarray testing [CMA]– but apparently not all. Since they do not (and cannot) assess that, this number is meaningless. The meaninglessness of this is amplified by the eligibility criterion of “cohort of patients 49 suspected of having a genetically caused rare disease”. There are no criteria for this and thus we cannot know what it means. It is capped by this ‘conclusion’: “yielded a particularly high proportion of ultra-rare diagnoses” what does “particularly high” mean and what is this being compared to?

**Response:** We thank the reviewer for these observations. The aims of the present study were to perform a systematic investigation of the generated phenotypic and molecular genetic data of patients who had undergone exome sequencing in order to determine the yield of ultra-rare diagnoses and of novel genotype-phenotype associations; and to determine whether the addition of machine learning and AI tools improved diagnostic effectiveness and efficiency for ultra-rare disorders.

We agree that the diagnostic yield of ES is dependent on the tests that were conducted prior to ES**.** However, evaluation of the effectiveness and efficiency of genetic testing is the basis for improved health economics, and thus represents an important contribution to public health research. The testing with NGS panels prior to ES explains why we established relatively few higher incidence diagnoses and more lower incidence diagnoses (which we refer to as ultra-rare). This is also shown in Supplemental Figure 5, in which we compare the molecular diagnoses established in the TRANSLATE NAMSE ES cohort with those established in a rare disease program in the UK. To clarify these issues, we have now amended the respective text.

**Reviewer #3:** No comparisons should be claimed unless statistical significance is assessed. “diagnostic yield was 5 percentage points higher for children (32%) than for adults (27%)” is one example.

**Response:** We thank the reviewer for this comment. For all comparisons, we now indicate the respective statistical tests and results. While we understand the reasoning of the reviewer, comparisons can be meaningful even in the absence of statistical significance. In this case, the data must be interpreted. The revised text reads: “The diagnostic yield was slightly higher in children (32%) than in adults (28%, p=0.13, Fisher’s exact test, Figure 2a) and two-fold higher in patients assigned to the category “neurodevelopmental disorder” than for all other disease categories (42% vs. 22%, p<0.001, Fisher’s exact test with Bonferroni correction; for single comparisons between disorder groups see Figure 2b)”

**Reviewer #3:** Why were some variants annotated by VEP and some by Jannovar?

**Response:** We thank the reviewer for this comment, and agree that this point must be clarified. The four data-generating sites used differing bioinformatic pipelines. For example, Berlin used Jannovar and VarFish, which is an open-source software tool kit that the Berlin team is in the process of developing (Holtgrewe et al., 2020; Jäger et al., 2014). In contrast, Tübingen established a pipeline that builds on VEP. To monitor the quality of these pipelines, ring trials were conducted between the four labs. These revealed excellent congruence.

**Reviewer #3:** The accuracy of the PEDIA top 10 is not valid unless the human assessment was blinded to the top 10 list. Nothing about blinding was stated in the methods.

**Response:** We are unsure whether we have understood the reviewer’s comment correctly, and apologize in advance if this is the case. The purpose of our work was not to compare performance between man and machine. Thus, there was no need to blind any human test participants. For all cases, the entire ES data has been evaluated. A case was referred to as “solved” if a molecular diagnosis could be established according to ACMG criteria. For this set the accuracy of PEDIA has been measured and compared to existing variant prioritization algorithms.

**Reviewer #3:** The ASXL3 result is perplexing. That the VAF was so low and low quality in the dad makes the call very suspicious. This has to be confirmed with deeper NGS sequencing, Sanger is just not adequate for this purpose. In the second case it is not valid to conclude mosaicism with a single read. The third case was found by simple pedigree analysis and not detected in the parent at all. This is all quite unimpressive. Again, they claim this is higher than previous work, but with no statistical support.

**Response:** We thank the reviewer for bringing this issue to our attention. In the revised manuscript, we have amended the formulation of the respective text in order to avoid the impression that we can draw definite conclusions. Instead, we explain that our aim was to present cases in which the entirety of the (sometimes indirect) evidence points to the existence of low-level parental mosaicism. The first case is indeed perplexing. On reflection, we agree with the reviewer that Sanger sequencing is insufficient in terms of excluding low-level paternal mosaicism. In response to the reviewer’s comment, we therefore re-sequenced both parents at a higher coverage, and now provide additional evidence for our conclusion.

In the second case, we also agree entirely with the reviewer that mosaicism cannot be unequivocally proven by a single maternal read. Our intention was to point out that maternal mosaicism was the probable explanation for this single read in combination with the presence of the corresponding variant in two sons. We have now reformulated the respective text for this case, as well as that for family 3.

We also agree with the reviewer that the numbers are too small for statistical considerations and due to a different study design also not comparable to the literature. We therefore refrain from an interpretation of potentially increased rates of parental mosaicism in our cohort and simply state this number due to its important clinical implications.

**Reviewer #3:** Their criteria for establishing novel gene-disease associations are simplistic. An entire Nature paper (PMID 24759409=MacArthur, Nature 2014) was written on this terribly difficult problem and it seems inappropriate for this group to suggest four simple bullet points to resolve this. The problem is that the mix of evidence is slightly different in every case. Reducing it to four criteria and a point score is naïve, at best.

On lines 270-272 they state “the concept of a clear separation between diagnostics and research cannot and should not be applied for patients with ultra-rare disorders.” This is a rather grand claim that appears to challenge the framework for research protections for human subjects. It is neither explained as to what they actually mean by this nor is it justified. One cannot simply discard human research ethics standards in this manner.

**Response:** We thank the reviewer for this important comment. We agree that the establishment of new gene-disease associations is highly complex and difficult to represent in terms of a scoring system. Furthermore, it was not our intention to propose a new score for that purpose. Instead, our evidence score was based on the established scoring systems, of MacArthur et. al. (2014), ClinGen (Strande et al., 2017), and Lieberwirth et al. (2022). These systems are now referenced. As proposed in these three systems, we first defined characteristics by which the level of evidence for the manually identified candidates could be assessed. These characteristics included gnomAD constraint metrics, inheritance, and functional data. Using these characteristics, we then calculated an evidence score. By applying our scoring system, we provide a detailed and objective record of all available evidence for each of the identified candidate genes. In our opinion, this information is of interest to the international rare disease community.

To clarify that only manually identified candidates were scored, we have amended the respective sentence in the online methods. This now reads: “As in the ClinGen approach and as suggested by others, we defined characteristics, including gnomAD constraint metrics, inheritance, and functional data, by which the level of evidence of the manually identified candidates could be assessed (Lieberwirth et al., 2022; MacArthur et al., 2014; Strande et al., 2017) (Supplemental Table 3).”

We also acknowledge that the wording in the original manuscript was inaccurate, and we agree with the reviewer that a separation between diagnostics and research cannot and should not be eliminated. We have now modified this statement accordingly. The revised text reads:

To acknowledge the complexity and difficulty of identifying novel disease genes, we have added the following sentence to the Discussion: “Establishing novel disease-gene associations or conducting functional analyses for the reclassification of variants of uncertain significance (VUS) are a time-consuming and highly complex endeavors (MacArthur et al., 2014). Hence, from the present logistical perspective, such analyses are easier to perform in a research context than within the routine diagnostic context of clinical practice. However, these findings are of crucial importance for affected individuals and their families. Thus from a teleological perspective, in some rare disease cases, boundaries separating diagnostics and research are somewhat blurred. Therefore in the tertiary, academic setting, collaboration between experts from diagnostics and research is highly relevant for patients with suspected ultra-rare diseases and a lack of definitive diagnostic findings”.

**Reviewer #3:** Lines 283-284 are in German instead of English.

**Response:** We thank the reviewer for pointing out this error. The German language text has now been deleted. The German text was a remnant of an internal discussion during manuscript preperation relating to the issue addressed in of the previous comment. The German phrase “wissensgenerierende Krankenversorgung” cannot be translated literally into English. We hope that the paraphrase is now comprehensible.

**Reviewer #3:** In lines 180-188 it becomes clear that several of these findings have already been published elsewhere. Thus, these findings are not novel and this part of the paper is just a summary of part of the overall program.

**Response:** During the course of the study 34 novel and 23 candidate genotype-phenotype associations were delineated. These findings are described in the results section Novel Diagnostic-grade genes (DGG) and candidates. A detailed list of all cases is given as Supplementary Table S3. In this table all previous publications are referenced if applicable. There are three scenarios that have to be differentiated:

A) DGG status was established by a peer-reviewed publication that included at least one case of this study.

B) A case of this study was diagnosed during the course of this study with a disease-causing mutation in a DGG, and the DGG status was established by another group.

C) There is high evidence for a disease-causing mutation in a novel gene.

A) and B) sum up to a total of 34. If DGG belongs to A) it is referenced in the main text (in total 13 references, which are references 20-32):

If a DGG belongs to 2) it is referenced in Supplementary Table 3 (in total 5 references):

C) adds up to 23 and require no references.

**Reviewer #3:** They highlight case 393 as a PEDIA success. But to me, it looks like a GS success – the finding of that de novo deletion would have led to the diagnosis. I don’t understand why they credit this to the gestalt software.

**Response:** We agree with the reviewer that the disease-causing 4.7kb deletion in this case would have been identified more easily with GS. However, as described in more detail in the revised manuscript, it would also have been possible to identify this deletion with Pindel in ES data (Brand et al., 2022). Nevertheless, due to the enrichment process, the quality of the CNV calls would be lower in ES than in GS, and the much higher rate of false positive calls would render data analysis more time-consuming. Thus, the PEDIA approach also facilitates the efficient analysis of this noisy data.

**Reviewer #3:** It is formally incorrect to say that “two brothers with the main symptom of intellectual disability carried a heterozygous (case 435), likely pathogenic variant in FOXG1” They are not carriers if they are affected. See PMID 27657676.

**Response:** We thank the reviewer for pointing out this error. We have now modified the text accordingly. The revised text reads: “In family 2, which included two siblings with the clinical feature of intellectual disability (case 435 and his brother), a heterozygous, likely pathogenic variant in *FOXG1* (*FOXG1* syndrome, OMIM #613454) was detected in both individuals.”

**Reviewer #3:** It is inappropriate to use slang in a scientific paper “Our findings highlight the importance of eyeballing (sic) parental NGS data for low-level mosaicism.”

We thank the reviewer for this comment. We have now deleted the term “eyeballing”, and replaced this with the phrase “visual inspection of”.
